# Early detection and improved genomic surveillance of SARS-CoV-2 variants from deep sequencing data

**DOI:** 10.1101/2021.12.14.21267810

**Authors:** Daniele Ramazzotti, Davide Maspero, Fabrizio Angaroni, Marco Antoniotti, Rocco Piazza, Alex Graudenzi

**Affiliations:** Dept. of Medicine and Surgery, Univ. of Milan-Bicocca, Milan; Dept. of Informatics, Systems and Communication, Univ. of Milan-Bicocca, Milan; Inst. of Molecular Bioimaging and Physiology, Consiglio Nazionale delle Ricerche (IBFM-CNR), Segrate, Milan; Bicocca Bioinformatics Biostatistics and Bioimaging Centre – B4, Milan

## Abstract

In the definition of fruitful strategies to contrast the worldwide diffusion of SARS-CoV-2, maximum efforts must be devoted to the early detection of dangerous variants. An effective help to this end is granted by the analysis of deep sequencing data of viral samples, which are typically discarded after the creation of consensus sequences. Indeed, only with deep sequencing data it is possible to identify intra-host low-frequency mutations, which are a direct footprint of mutational processes that may eventually lead to the origination of functionally advantageous variants. Accordingly, a timely and statistically robust identification of such mutations might inform political decision-making with significant anticipation with respect to standard analyses based on consensus sequences.

To support our claim, we here present the largest study to date of SARS-CoV-2 deep sequencing data, which involves 220,788 high quality samples, collected over 20 months from 137 distinct studies. Importantly, we show that a relevant number of spike and nucleocapsid mutations of interest associated to the most circulating variants, including Beta, Delta and Omicron, might have been intercepted several months in advance, possibly leading to different public-health decisions. In addition, we show that a refined genomic surveillance system involving high- and low-frequency mutations might allow one to pin-point possibly dangerous emerging mutation patterns, providing a data-driven automated support to epidemiologists and virologists.

## Introduction

The continuous evolution of the COVID-19 pandemic at the global scale has proven that pivotal efforts must be devoted by the scientific community to the timely identification and quantification of hazardous variants, i.e., those showing increased virulence, pathogenesis or ability to escape therapeutic strategies such as vaccines (*1, 2*). To this end, institutions such as the World Health Organization (WHO), the European Centre for Disease Prevention and Control (ECDC), the Centers for Disease Control and Prevention (CDC), and others are repeatedly updating the lists of the so-called variants of interests (VOIs), variants of concern (VOCs), variants under monitoring (VUMs) (also named Variants Being Monitored -VBMs) and de-escalated variants (DEVs) (*3–5*). Each variant is typically characterized by a set of spike mutations of interest (SMoIs) and is categorized according to different institution-specific molecular and epidemiological criteria (*6*). VOCs are usually associated with evidence of diminished effectiveness of treatments, increased transmissibility, immune escape and/or diagnostic escape. VOIs present genetic changes that are predicted or known to cause the same effect of a VOC, but have a limited prevalence (e.g., in circumscribed outbreak clusters). VUMs bear genetic markers sus-pected to impact the epidemic dynamics, but circulate at a very low level. Finally, a variant is classified as DEV if it is no longer circulating, or if there is solid evidence that it does not affect the overall epidemiological situation. In Table 1, one can find the list of SARS-CoV-2 variants listed as VOI, VOC, VUM or DEV by at least one of the three public health bodies as of October 26th 2021, and the related WHO label (in Greek letters), Pango lineage (*7*) and set of SMoIs (for further details on the categorization criteria please refer to the institution websites; note also that the Omicron variant was added to the list, despite being designed as VOC by the WHO on November 26th 2021).

**Table 1:**
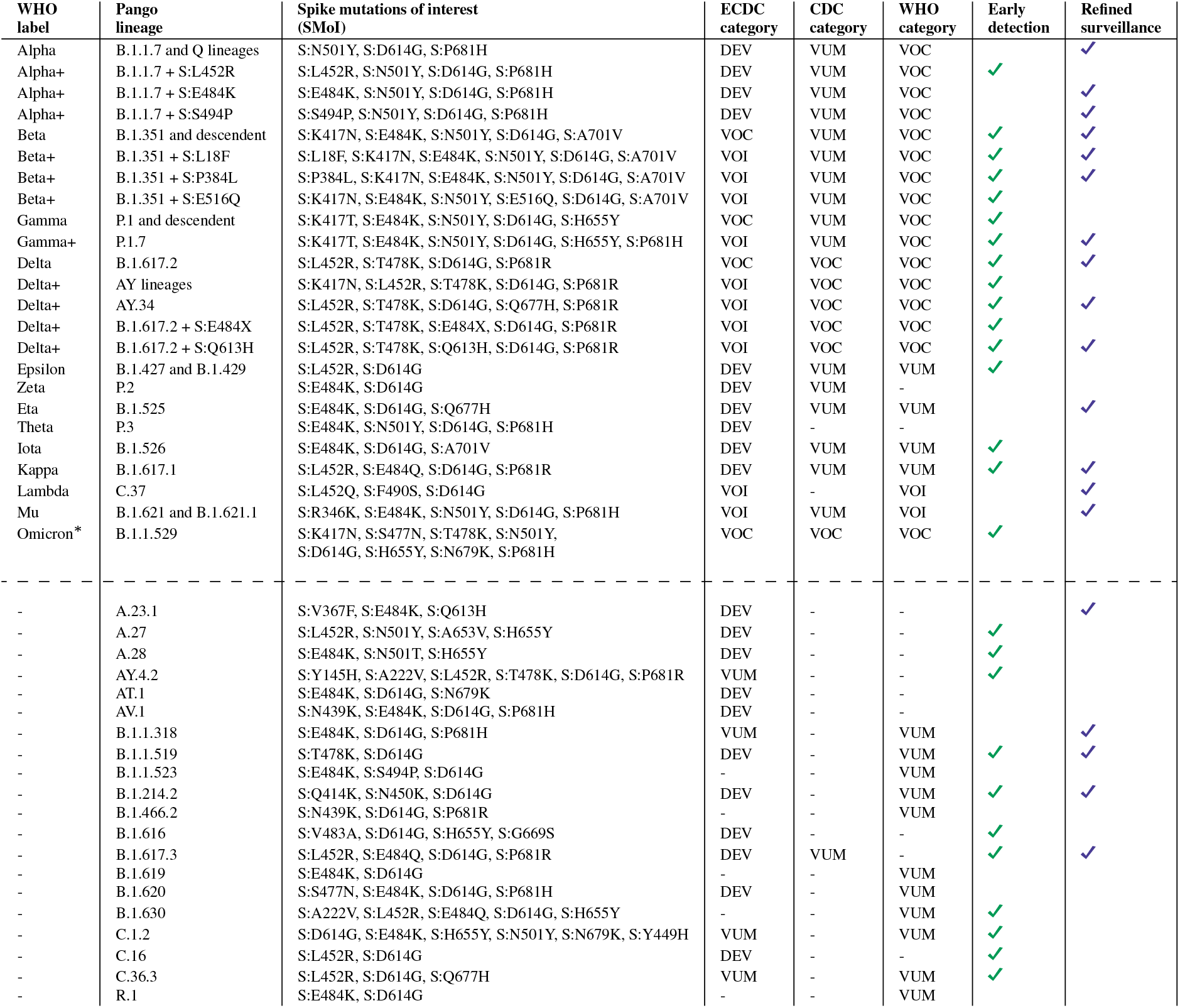
Hazardous SARS-CoV-2 variants. List of SARS-CoV-2 variants of concern (VOC), of interest (VOI), under monitoring (VUM), and de-escalated variants (DEV), updated on October 26th, 2021, as from the categorization of (*3–5*). Omicron variant was added to the list even if designated as VOC on November 26th. Information on the WHO label, the constituting Pango lineages (*7*), the associated spike mutations of interest (SMoI), and the institution-specific categories are shown. Variant labels marked with “+” include additional SMoIs with respect to the related upstream variant. In the last two columns, we report the variants for which either an early detection of the related SMoIs and/or a refined surveillance (via homoplasy analysis) is granted by exploiting deep sequencing data (see Results). Notice that A.23.1 and B.1.525 (Eta) are included in the list of so-called variants of note in the Cov-Lineages.org lineage report (*8*). *Notice that, currently, no SMoIs are associated to the Omicron variant. Here, we indicate the S mutations present in such variant *and* identified as SMoI in at least one of the remaining 43 variants.

In the last couple of years, the analysis and surveillance of SARS-CoV-2 variants has bene-fited from the surge of sequencing data that are first generated and then made available on portals such as GISAID (*9*), Nextstrain (*10*) or Cov-Lineages.org (*8*). However, the large majority of available datasets include *consensus sequences* generated (with distinct criteria) from whole-genome sequencing experiments performed on primary isolates, rather than deep sequencing data. In fact, the proportion of deep sequencing datasets shared on public repositories has been significantly lower than that of consensus sequence during the course of the pandemic (see Figure 1). Even if this state of affairs is progressively evolving, as of August 2021, 4,558,675 consensus sequences are available on the GISAID database (*9*), whereas only 975,767 samples (≈ 21.4%) are included in (Illumina paired-end) sequencing datasets accessible (from different sources) on the NCBI website (*11*), of which only approximately 50% are of high-quality (see Methods for further details). Furthermore, no unique database exists including the different datasets at hand, whose download and processing require a significant amount of manual curation and computation time.

**Figure 1:**
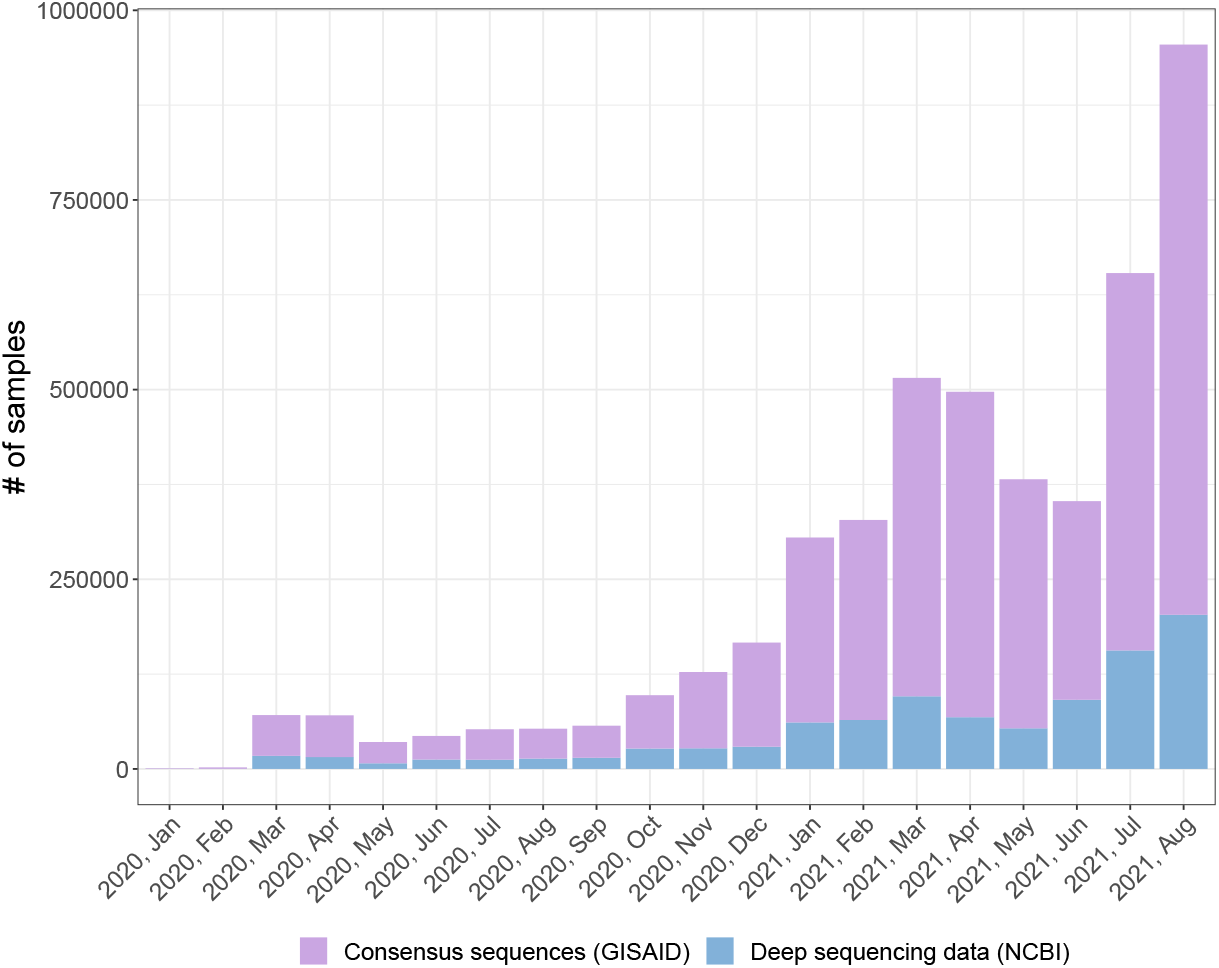
SARS-CoV-2 samples in public repositories. Number of SARS-CoV-2 samples for which either deep sequencing data or consensus sequences are available, grouped by month in which the related dataset is released in the period January 2020 – August 2021. Source databases are NCBI (*11*) for deep sequencing data and GISAID (*9*) for consensus sequences (update: August 2021).

Deep sequencing data are indeed essential to characterize the intra-host mutational land-scape of viral samples and, especially, to identify the presence of minor mutations, i.e., single-nucleotide variants (SNVs) and indels detected with *low mutation frequency* (MF, which – roughly – is the ratio of alternative allele reads over the total reads for a specific genome position). Minor mutations are – by definition – not included in consensus sequences, but characterize the heterogeneous ensemble of viral subpopulations, known as *quasispecies* (*12, 13*), that are typically present in single-hosts. Most important, every new genomic mutation first originates as minor within single hosts, due to replication errors of viral polymerases that are often induced by host-related mutational processes (*14, 15*).

*Minor mutations* are generally purified, due to detrimental effects on the fitness of the virus, and are significantly affected by transmission events (e.g., bottlenecks), which hamper their diffusion in the population (*16*), as opposed to *fixed mutation*, which are usually transmitted from a host to another during infections. However, certain minor mutations may provide the virus with a fitness advantage, for instance in terms of enhanced reproductive potential or increased infectiveness and, accordingly, are positively selected, first within hosts and, later, in the population of infected people. For these and many additional transmission-related phenomena, the MF of certain minor mutations can sometimes increase both within single hosts and across infection chains. Once the MF exceeds a certain threshold (typically around ∼ 50%), the mutation fixates, meaning that the related viral subpopulation has become dominant within a given host. Only then the mutation is included into consensus sequences. Among the set of fixed mutations, some (e.g., the SMoIs) will eventually contribute to the origination of hazardous variants, subsequently impacting the course of the epidemic.

A finer characterization of the minor mutation landscape is therefore beneficial to: (*i*) intercept impactful mutations prior to their fixation, (*ii*) assess the presence of dangerous minor mutations in samples exhibiting circulating variants. Both aspects are essential for the definition of an effective genomics-informed epidemiological surveillance system, which may drive the design of timely public-health interventions, with substantial repercussions in terms of epidemiological dynamics and socioeconomic costs (*17–20*). In this regard, note that the characterization of the minor mutation landscape has already been successfully exploited to investigate drug-resistance, contagion chains, bottleneck effects and mutational signatures, for a number of infectious diseases including COVID-19 (*13, 14, 21, 22*).

In support of our claims, here we provide the largest up-to-date worldwide study of deep sequencing data of SARS-CoV-2 samples, which includes 220, 788 high-quality samples from 137 distinct datasets. Our analyses are first focused on a list of 44 variants and related 35 SMoIs, included in the lists of hazardous variants by the WHO, the ECDC and the CDC. We also analyse a list of 95 further spike mutations that have not currently been associated to any known variant, but display significant diffusion patterns. Since attention was recently raised on the functional role of mutations hitting the nucleocapsid (N) protein (via the analysis of SARSCoV-2 virus-like particles (*23*)), we also investigated a list of 13 N mutations with potential functional effect (here labelled as *N mutations of interest*, NMoIs), and a further list of 82 highly diffused N mutations.

In brief: (*i*) we prove that the identification of several S and N mutations could be anticipated of a significant time-span with respect to standard analyses based on consensus sequences, and (*ii*) we highlight that a significant number of samples harboring circulating variants display homoplastic minor mutations, which might lead to the origination of new variants.

## Results

The analyses presented in this work are focused on: (*i*) the set of 35 S mutations (SMoIs) associated to the 44 SARS-CoV-2 variants included in the lists of VOCs, VOIs, VUMs or DEVs in at least one of the WHO, the CDC and the ECDC websites, as of October 26th 2021 (see Table 1; Omicron variant was labeled as VOC on November 26th 2021), (*ii*) a list of additional 95 S mutations significantly diffused in the population, (*iii*) a list of 13 N mutations with potential functional effect identified in (*23*) (NMoIs), (*iv*) a further list of highly diffused 82 N mutations. No considerations on the molecular properties or the epidemiological features of such mutations/variants are purposely reported, for which the readers are referred to the related literature.

### Early detection of mutations of interest

#### Spike mutations of interest associated to known variants (SMoIs)

One of the major differences in employing deep sequencing data instead of consensus sequences lies in the possibility of detecting, in principle, any genomic mutation with great temporal advance. To corroborate this claim, we first analyzed separately each SMoI associated to the hazardous variants included in Table 1. A total amount of 35 distinct mutations were analysed, involved in 44 variants (any variant can include one or more Pango lineages (*7*)).

In particular, we computed: (*i*) the distribution of the MF of all the SMoIs on all samples, (*ii*) the prevalence of each SMoI in the population when either detected as minor (MF ≥ 5% and < 50%) or fixed (MF ≥ 50%), with respect to collection date (grouped by month) and location (grouped by continent).

Overall, 4 (out of 35) SMoIs were detected as minor (in at least 5 samples) at least one month prior to their initial detection as fixed at the global level, i.e., S:Q414K in March 2020, S:L452R in September 2020, and S:H655Y in January 2021, and 2 additional SMoIs in at least one of the considered geographical regions, i.e., S:L18F, S:T478K and S:A701V, in September 2020 in Africa (see Figure 2). The 6 SMoIs detected in advance characterize a large number of hazardous variants (see Table 1) and, in particular: S:Q414K is associated to one variant^1^, S:L452R to 14 variants^2^, S:H655Y to 8 variants^3^, S:L18F to one variant^4^, S:T478K to 8 variants^5^ and S:A701V to 5 variants^6^.

**Figure 2:**
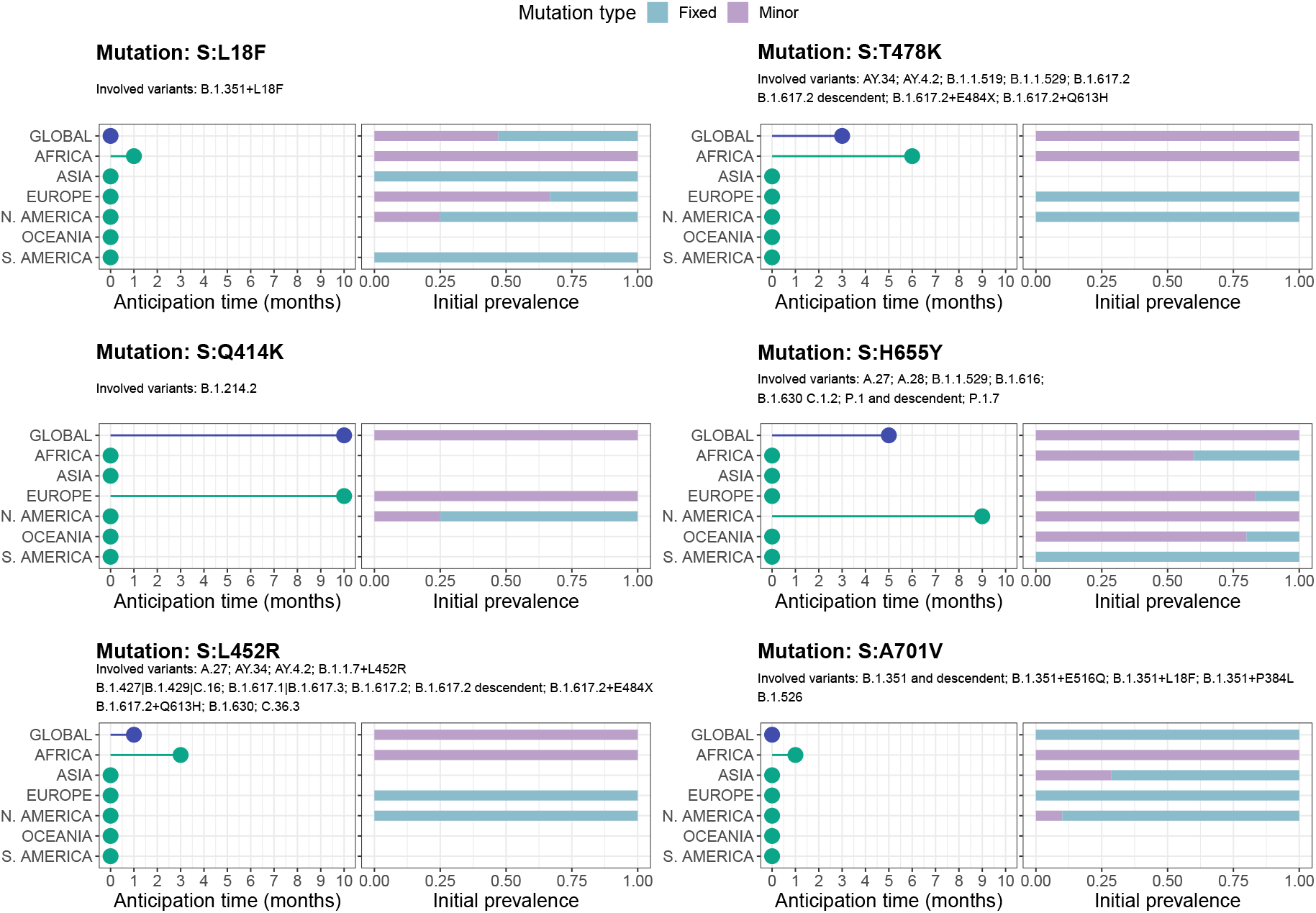
Early detection of 6 SMoIs associated to hazardous variants from deep sequencing data. Analysis of SMoIs: S:L18F, S:Q414K, S:L452R, S:T478K, S:H655Y and S:A701V (see Table 1). The leftmost panels report how many months in advance any SMoI was detected as minor (MF ≥ 5% and < 50%) in at least 5 samples, while been undetected as fixed (MF ≥ 50%), either at the global scale or in any of the 6 considered geographical regions. The rightmost panels show the proportion of samples exhibiting the mutation either as minor or fixed, in the first month in which the mutation was firstly detected in at least 5 samples, either at the global or the local scale. For each SMoI the related variants are also displayed.

Moreover, 4 additional SMoIs were initially detected *both* as minor (in at least 5 samples) and fixed (in at least 1 sample) at the global scale (S:V367F, S:K417N, S:A653V, S:Q677H), and another SMoI in at least one geographical region (S:D614G in North America), demonstrating that the circulation of SMoIs can be underestimated by considering consensus sequences only.

In Figure 3, one can find the in-depth analysis of mutations S:L452R and S:H655Y, whereas the analysis of all remaining early-detected S and N mutations is presented in Supplementary Figures 3–36. More in detail, at the global scale, mutation S:L452R is first observed as minor in Africa in September 2020 and as fixed in North America in October 2020 (1 month in advance). The anticipation is remarkably amplified when considering the local scale. In fact, in Africa the mutation is observed as fixed only in December 2020, that is 3 months later.

**Figure 3:**
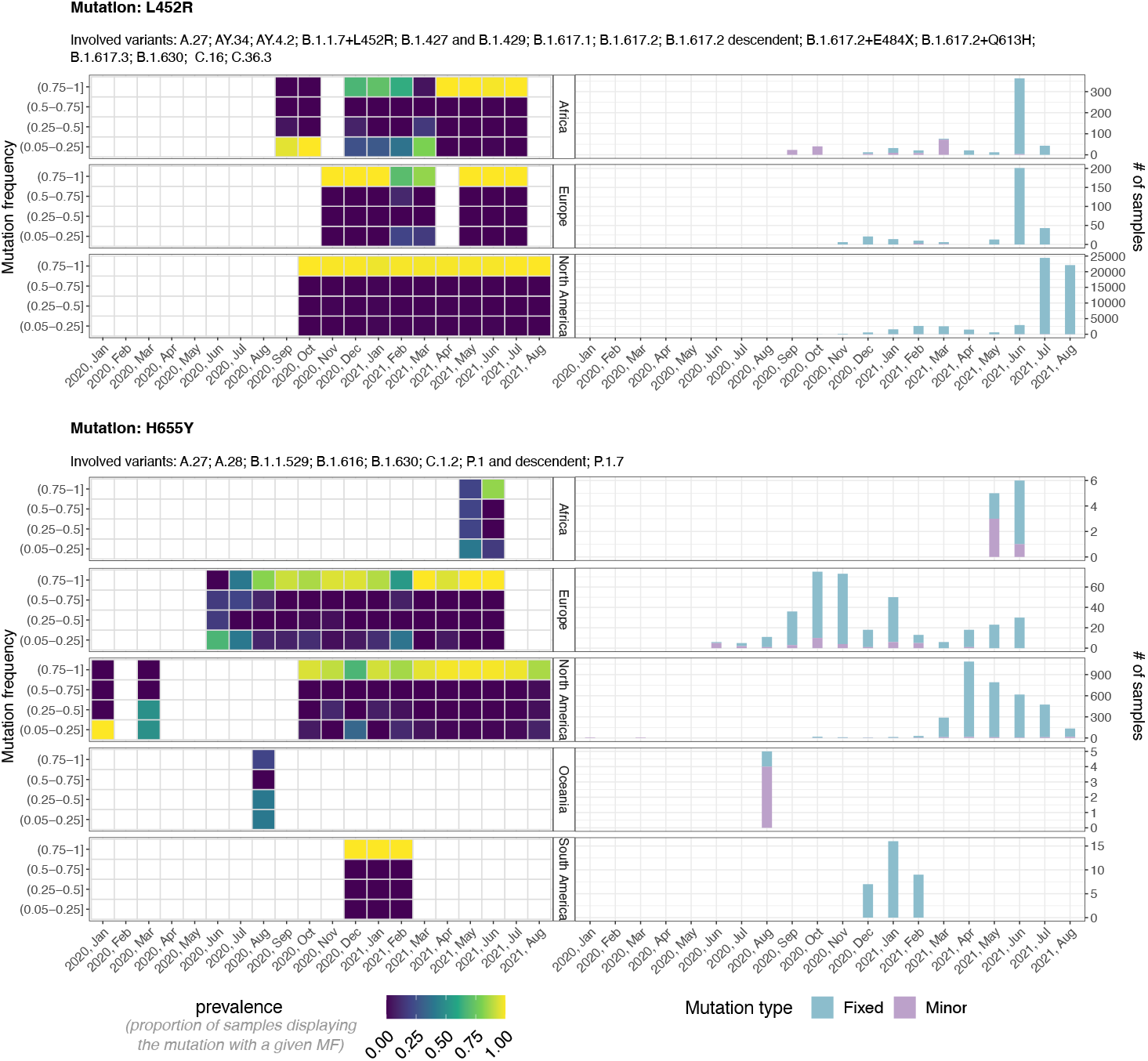
Mutant frequency and prevalence variation in time of SMoIs S:L452R and S:H655Y. The leftmost panels return the distribution of the mutation frequency (MF) of all samples with SMoIs S:L452R (upper panels) and S:H655Y (lower), grouped by month and geographical region. Each cell shows the proportion of samples showing the mutation with that specific MF. The rightmost panels show the number of samples showing the mutations either as minor (MF ≥ 5% and < 50%) or fixed (MF ≥ 50%). The lineages associated to both variants are also displayed.

A similar trend is observed for mutation S:H655Y, which is firstly detected as minor in January 2020 in North America, and as fixed in June 2020 in Europe (5 months in advance at the global scale) and in October 2020 in North America (9 months in advance at the local scale). Notice also that when mutation S:H655Y was first detected in Europe, Africa and Oceania, the large majority of samples exhibited it as minor. These important results show that, once a variant is identified as VUM, VOI or VOC, it is possible to detect the related mutations considerably before their fixation at both the global and the local scale, allowing for a timely implementation of containment strategies.

As an aside note, the distribution of the MF proves that most SMoIs are present either at a very low or at a very high frequency within hosts, suggesting the presence of strong purifying selection and of bottlenecks, as already observed in (*14*) (see Supplementary Figures 3–36).

#### Additional spike mutations

Even if a mutation is not associated to any of the known variants, it is possible to investigate its selection and fixation dynamics with remarkable anticipation by looking at its MF variation in time and its diffusion in the population.

To this end, we analysed the list of (single-nucleotide) mutations meeting the following criteria: (*i*) falling on the spike gene, (*ii*) not being associated to any of the variants of Table 1, (*iii*) detected (with MF *>* 5%) in at least 50 samples in the whole considered period, (*iv*) detected (with MF *>* 5%) in at least 1% of the samples in the month in which detected with the highest prevalence. The final list includes 95 spike mutations (S:N30H was excluded from the analysis after manual curation).

In brief, 6 (out of 95) mutations were initially found as minor at the global scale (S:W152C, S:S297L, S:C361S, S:G446V, S:A570D, S:T791K) (See Figure 4), and 11 additional mutations at the local scale (S:T95I, S:T167I, S:R682W, S:R685L, S:R685S, S:T716I, S:T791I, S:A892V, S:D1118H, S:G1124V, S:W1214G) (shown in Supplementary Figure 1 for reasons of space). Interestingly, mutation S:T95I is associated to Omicron variant, while currently not being listed as SMoI.

**Figure 4:**
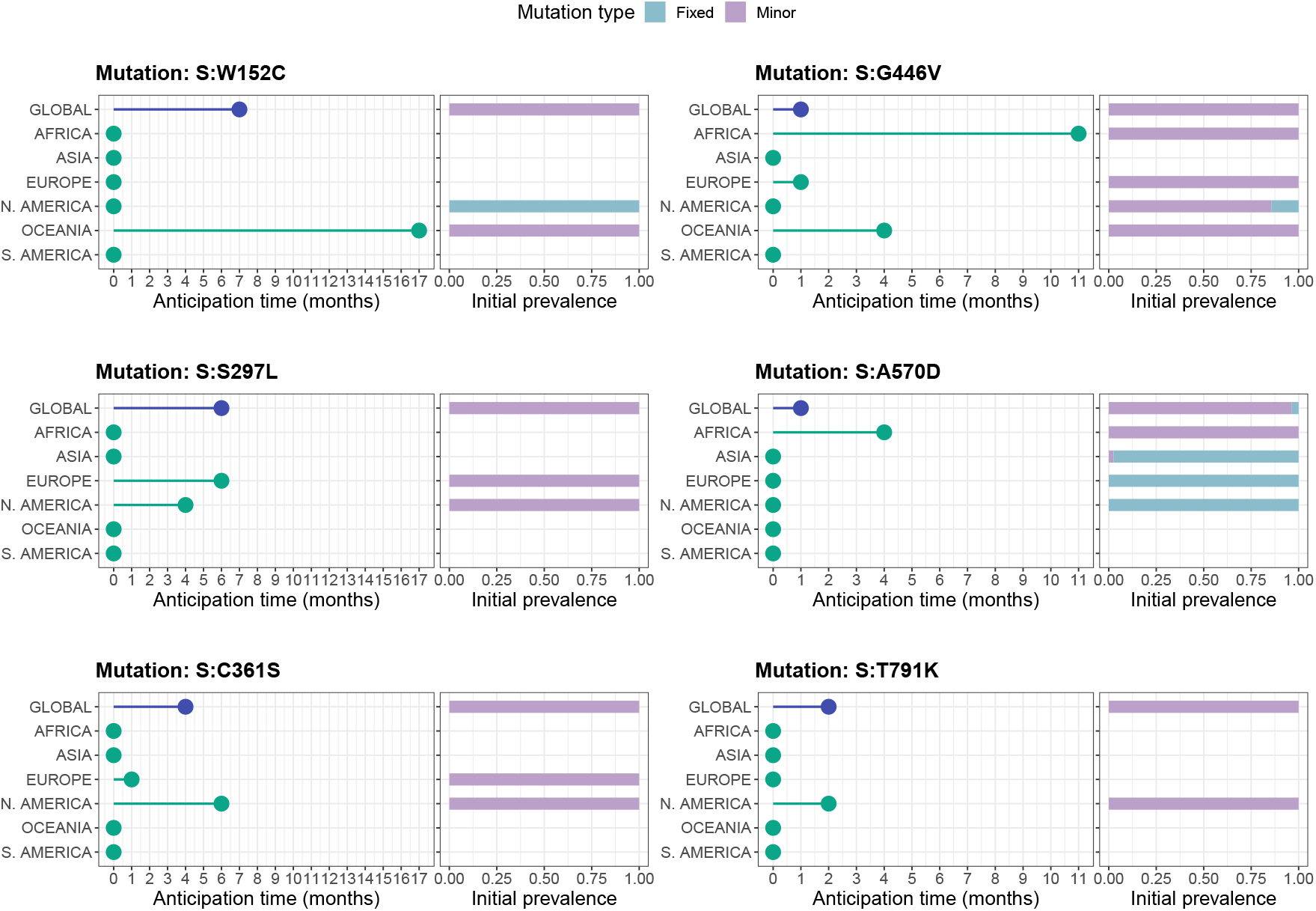
Early detection of 6 S mutations not associated to known variants. Analysis of 6 S mutations originally detected as minor (in at least 5 samples) and only successively as fixed at the global scale, namely: S:W152C, S:S297L, S:C361S, S:G446V, S:A570D, S:T791K. For further details please refer to the caption of Figure 2. S mutations first detected as minor at the local scale are shown in Supplementary Figure 1.

Also in this case, a significant anticipation is granted by the analysis of deep sequencing data, up to 7 months at the global and up to 17 months at the local scale, respectively.

Among the many mutations, S:V1264L was recently associated to Delta-1 variant and was hypothesized to underlie the outbreaks in Indonesia, Singapore and Malaysia (*24*). Mutation S:W152C is associated to the lineage B.1.429, but not included in the list of SMoIs from any of the considered institutions. S:W1214G was identified as destabilizing in (*25*). These further results proves that the analysis of the minor mutation landscape might be effective to intercept hazardous S mutations prior to their fixation in the population, even when not associated to lists of known variants.

We finally note that, 9 additional highly diffused S mutations were initially detected *both* as minor (in at least 5 samples) and fixed (in at least 1 samples) at the global scale (S:Q23H, S:A27S, S:S98F, S:F157L, S:L176F, S:K529M, S:T547I, S:G769V, S:V1264L), and other 6 S mutations in at least one geographical region (S:L5F, S:D80A, S:T95I, S:G446V, S:A892V, S:G1124V), demonstrating that relevant information might be missed by looking at consensus sequences only.

Mutation S:G446V shows particularly interesting dynamics as it has been observed mostly (*>* 90% samples) as minor mutation since March 2020, but has been showing an increase in MF since November 2020, and is now observed as a fixed variants approximately in 50% of the samples presenting the mutation in July 2021 and August 2021. Moreover, mutation S:G446V has been associated to attenuate monoclonal and serum antibody neutralization (*26*).

#### Nucleocapsid mutations

We repeated the analysis by first focusing on the list of 13 NMoIs selected in (*23*). 4 of them, in particular, were associated to a significant increased mRNA delivery and expression from the analysis of SARS-CoV-2 virus-like particles (N:P199L, N:S202R, N:R203K and N:R203M), and it was also hypothesized that one of such mutation may be responsible for the increased spread of variants including Delta (N:R203M). Furthermore, we selected an additional list of 83 highly diffused N mutations, with the criteria employed for the additional S mutations and described above (mutations N:D3E, N:D3H, N:D3V and N:K256* were removed from the analysis after manual curation).

As a result, 3 (out of 96) N mutations were initially found only as minor (in at least 5 samples) at the global scale (N:L219F, N:A254S, N:A254V), and 7 further mutations at the local scale (N:H145Y, N:S197L, N:G204A, N:L222M, N:Q244K, N:A305V, N:K374N), with distinct anticipation according to the cases, up to a maximum of 19 in the most extreme case (see Figure 4).

Of such mutations, N:D377Y was identified as NMoI in (*23*), and in Africa was discovered as minor 3 months in advance. In Figure 5, one can find such mutations (N:D377Y), in addition to the three mutations first detected as minor at the global scale (N:L219F, N:A254S, N:A254V), whereas the remaining mutations are shown in Supplementary Figure 2.

**Figure 5:**
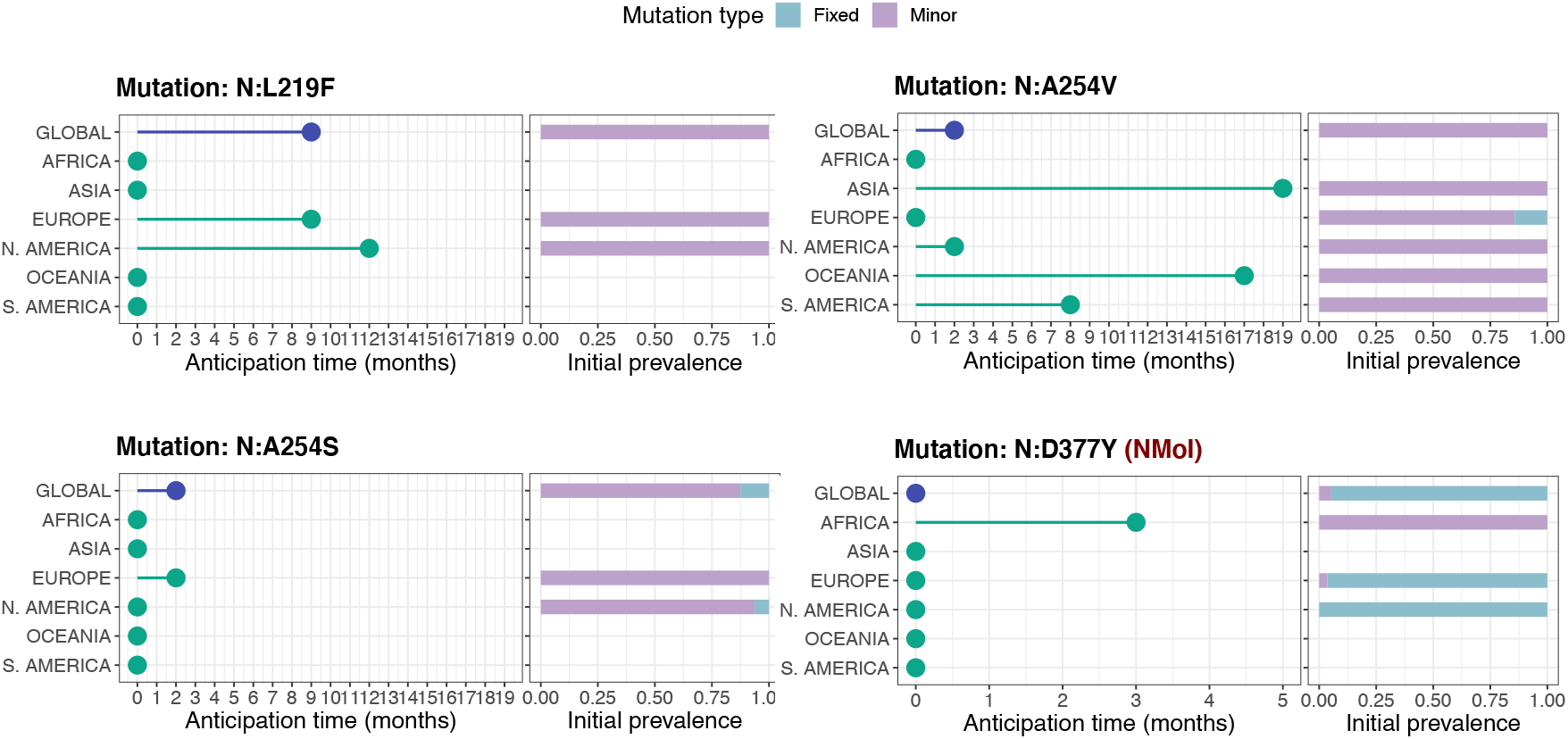
Early detection of N mutations. Analysis of NMoI N:D377Y, and of the three highly diffused *N* mutations originally detected as minor (in at least 5 samples) and only successively as fixed at the global scale, namely: N:L219F, N:A254S, N:A254V. For further details please refer to the caption of Figure 2. N mutations first detected as minor at the local scale are shown in Supplementary Figure 2.

Although ad-hoc investigations on the possible functional effect of such mutations are clearly required, these findings demonstrate the effectiveness of deep sequencing data analyses to intercept possibly hazardous mutations.

In further support to this claim, let us also notice that 1 additional NMoI was initially detected *both* as minor (in at least 5 samples) and fixed (in at least 1 samples) (N:M234I), and 8 highly diffused N mutation (N:P13T, N:H145Y, N:R185C, N:P168S, N:G238C, N:E253D, N:S327L, N:D415G) at the global scale.

### Improved genomic surveillance of circulating variants

In addition to the early detection of mutations, deep sequencing data are important for the characterization of the intra-host diversity of SARS-CoV-2 samples that are already associated to circulating variants, overcoming the intrinsic limitations of studies on consensus sequences. This analysis has important repercussion in terms of genomic surveillance.

In fact, homoplastic minor mutations (i.e., retrieved in distant lineages, with sufficient sample size) are typically not related to transmission events, but emerge independently in unrelated hosts, either due to a possible fitness advantage or to mutational hotspots (*22*). Accordingly, they should be flagged and carefully considered, as they might possibly lead to the origination of new dangerous variants, if positively selected due to any underlying functional advantage. Thus, their characterization might allow one to design opportune alert systems and timely intervention strategies.

We considered the list of mutations previously analyzed (35 SMoIs, 95 additional S mutations, 13 NMoIs and 83 additional N mutations), and assessed their presence in the minor state (MF ≥ 5% and < 50%) in the samples associated via Pangolin (*27*) to the variants included in Table 1 (samples assigned to the “Other” category were excluded from the analysis, whereas, currently, Pangolin does not associate samples to the Omicron variant). In Figure 6, one can find the prevalence of the selected minor mutations in the samples associated to the different variants. Only the mutations retrieved in at least 1% of the samples in at least one variant are considered.

**Figure 6:**
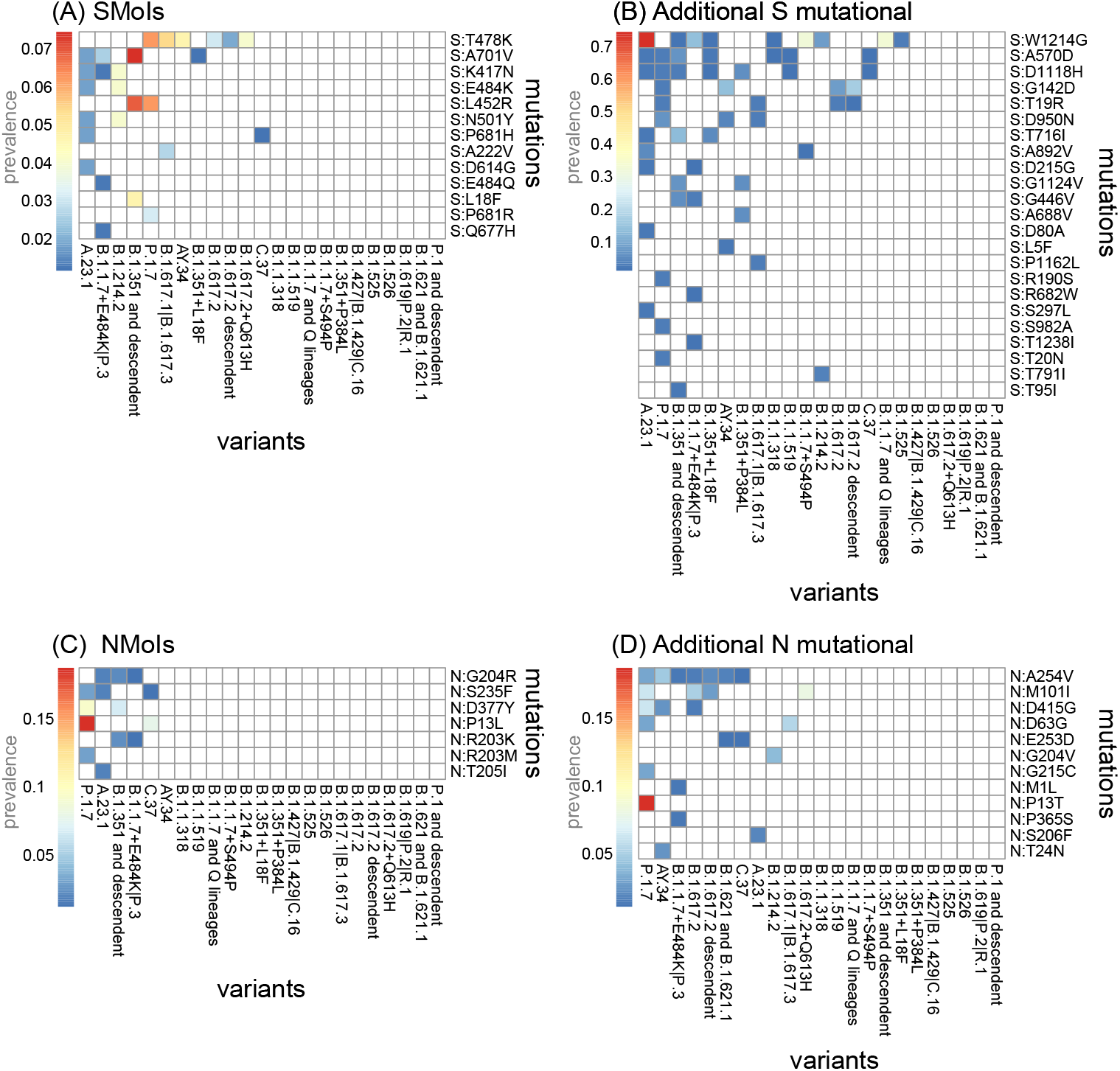
Analysis of homplastic minor variants. The heat-maps show the prevalence (i.e., number of samples over the total) of the SMoIs (panel A), additional highly-diffused S mutations (B), the NMoIs (C) and the additional highly-diffused N mutations (D) retrieved as minor (MF *>* 5% and ≤50%) in the samples associated to the variants of Table 1 via Pangolin (*27*). Only the mutations observed in at least 1% of the samples of any variant are shown.

In detail, 13 (out of 35) SMoIs are retrieved as minor and homoplastic (in at least 1% of the samples) and, in particular, 7 SMoIs are observed in more than one additional variant, namely S:T478K (in 6 variants), S:A701V (in 4), S:K417N (in 3), S:E484K (in 2), S:L452R (in 2), S:N501Y (in 2) and S:P681H (in 2). 23 (out of 95) additional S mutations were found as homoplastic, with the most notable examples being mutations S:W1214G, S:A570D, S:D1118H, S:G142D and S:T19R, which were found in at least 4 additional variants.

Similarly, 7 (out of 13) NMoIs were found as minor and homoplastic, with mutations N:G204R, N:S235F, N:D377Y, N:P13L and N:R203K in at least 2 variants. Finally, 12 (out of 83) additional N mutations were detected in multiple variants, with mutations N:A254V, N:M101I, N:D415G, N:D63G and N:E253D in at least 2 additional variants. This result points at a possible ongoing selection processes, supporting the hypothesis of an important, yet underestimated, functional impact of mutations of this protein in SARS-CoV-2 evolution.

Overall, 19 (out of 44) variants listed in Table 1 (dark purple check marks) display homoplastic minor (S or N) mutations of interest. This result confirms the benefits of analyzing deep sequencing data to pinpoint the emergence and positive selection processes of hazardous mutations, also in samples harboring known variants, with an improved resolution with respect to standard analyses of consensus sequences.

### Limitations

Any analysis of intra-host viral diversity highly depends on the quality of upstream variant/ haplotype calling, which is in turn closely related to the adopted technology and the testing criteria (*28*). When dealing with low-frequency mutations, one of the main criticisms lies in the difficulty of identifying true mutations from sequencing artifacts or phantom mutations, due, e.g., to mutational hotspots (*29*), as well as the possibility of dropouts, due, e.g., to uneven coverage (*30*). The analysis of distinct sets of minor variants might deceive statistical inference approaches leading to partially incorrect results, and this might apply to the case of early detection of mutations as well. To this end, many variant callers currently exist to correct for data-specific errors (*13*), and might be tested to assess the robustness of the results discussed hereby.

The limitations related to partial and inhomogeneous testing/sampling of the population are discussed in the next section.

## Discussion

Thanks to the largest-up-to-date analysis of deep sequencing datasets of SARS-CoV-2 samples, we proved that standard studies based on consensus sequences might be scarcely effective for the early detection of mutations of interests and for the fine monitoring of homoplastic variants that might lead to the origination of new variants. These aspects are even more relevant when considering the exceptional proportion of the COVID-19 pandemic and should be wisely considered in the ominous prospective of future epidemics.

Accordingly, a refined estimation of key epidemiological parameters (e.g., *R*_*t*_) from deep sequencing data might lead to significant differences in the predictions delivered by the wide range of currently available epidemiological models (*31, 32*), as well as in the accuracy and robustness of phylodynamics methods, which in most cases rely on consensus sequences (*33*). This might affect, in turn, the geo-temporal narrative on variants origination, as well as that of infection chains and (multiple) introductions (*34*), possibly guiding improved testing and response strategies.

In this work, we started the analysis from the set of mutations related to known variants. Yet, a large set of additional S and N mutations were detected as minor with great advance with respect to standard analyses. It would be important to combine this data-driven result with automated approaches aimed at identifying dangerous mutations/variants, e.g., via molecular simulations of the functional effects of genomic changes, as the binding energy between spike proteins and ACE2 or the binding affinities to antibodies (*35–39*).

Furthermore, the difference in the anticipation window related to the distinct geographical region suggests a straightforward way of improving current surveillance practices: once a variant start being monitored, even if already fixed in some areas, the analysis of the related minor mutations on different locations might allow one to intercept outbreak clusters with great advance, as well as to better estimate its overall prevalence.

It is also vital to point out that both the overall number of (minor) mutations detected in advance, and the magnitude of anticipation, would have dramatically benefited from the possibility of accessing a larger number of deep sequencing datasets, especially at the beginning of the epidemic. This issue mostly resulted from the absence of shared standards for testing and sequencing, which also contributed to the origination of relevant sampling biases and geographical inhomogeneities (*40*).

In this respect, and given the strong evidences of our results, a methodological paradigm shift seems to be essential and might take advantage of the ever-increasing computational power and of the pervasive data sharing networks currently available to the scientific community for the analysis of deep sequencing data of viral samples. We therefore advocate a general effort for the definition of a unique, standardized, shared, open, online, naturally FAIR-compliant (*41*) repository in which sharing (possibly pre-processed) raw sequencing data of viral samples.

## Materials and Methods

### Datasets

We analyzed a total 391,173 samples from distinct individuals obtained from 137 public NCBI BioProjects (see Supplementary Table 1 for the list of BioProjects and samples metadata). All selected FASTQ samples were paired-end amplicon sequencing data prepared following the COVID-19 ARTIC v3 Illumina library construction and sequencing protocol.

### Mutation calling

Mutation calling was performed by employing the iVar (version 1.3.1) (*42*) recommended pipeline to analyze SARS-CoV-2 ARTIC v3 amplicon sequencing data. In particular, we performed the following steps: 1) FASTQ files were mapped to the reference genome SARS-CoV-2-ANC (*14, 22*) with *bwa mem* (version 0.7.17-r1188) (*43*). 2) Sorted BAM files were generated from *bwa mem* results using SAMtools (version 1.10) (*44*). 3) ARTIC v3 primer sequences were trimmed using *ivar trim* command. 4) Trimmed sorted BAM files were built and indexed with SAMtools. 5) Mutation calling was performed from trimmed sorted BAM files using *ivar variants*. 6) Finally, *samtools depth* was used to extract coverage information from trimmed sorted BAM files.

### Quality control

Quality control was performed on the mutations obtained with *iVar variants*. We first selected (ultra) deep sequencing samples with a coverage of at least 100 reads in at least 90% of viral genome. Then, we perform further filtering by selecting only mutations with variant frequency of at least 5%, coverage of at least 100 and p-value resulting from *ivar variants* less than 0.01. Finally, samples with more than 100 mutations (after filtering) were remove, to obtain a final dataset comprising a total 220,788 samples and 7,855,379 (88,889 unique) mutations (see Supplementary Table 1).

### Amino acid sequence annotation

We considered all nonsynonymous substitutions (i.e., we kept only single base mutations) and annotated them to the related amino acid sequence. To avoid ambiguities, we removed mutations spanning mismatching positions between SARS-CoV-2-ANC (*14, 22*) and other proposed SARS-CoV-2 reference genomes (*45, 46*), namely positions 8782, 28144 and 29095 of SARS-CoV-2-ANC were removed. This led us to a total of 4,962,209 and 46,903 unique amino acid changes (see Supplementary Table 2).

### Pangolin analysis

We created consensus sequences as input to Pangolin (*27*) from the mutations obtained from deep sequencing data as explained in *Mutation calling*. We considered mutations with MF ≥ 0.50, i.e., the standard consensus sequences as uploaded, e.g., on GISAID (*9*). We created consensus sequences for each sample by adding to the reference genome SARS-CoV-2-ANC (*14, 22*) sequence, the mutations insertions and deletions observed in the sample for each position and by choosing the one at higher MF if multiple mutations were detected in the same position. On such inputs, Pangolin was executed with standard parameters.

## Data Availability

All data analyzed in this study are available online at the National Center for Biotechnology Information (NCBI) or the European Nucleotide Archive (ENA).

## Acknowledgments

This work was partially supported by the Elixir Italian Chapter and the SysBioNet project, a Ministero dell’Istruzione, dell’Universitàe della Ricerca initiative for the Italian Roadmap of European Strategy Forum on Research Infrastructures and by the Associazione Italiana per la Ricerca sul Cancro (AIRC)-IG grant 22082. DR and FA were partially supported by a Bicocca 2020 Starting Grant. DR was also supported by a Premio Giovani Talenti of the University of Milan-Bicocca. We thank Chiara Damiani, Lucrezia Patruno, Francesco Craighero and Gianluca Ascolani and for helpful discussions.

## Footnotes

1 B.1.214.2

2 B.1.1.7 + L452R (Alpha+), B.1.617.2 (Delta), AY lineages (Delta+), AY.34 (Delta+), B.1.617.2 + E484X (Delta+), B.1.617.2 + Q613H (Delta+), B.1.427 and B.1.429 (Epsilon), B.1.617.1 (Kappa), A.27, AY.4.2, B.1.617.3, B.1.630, C.16, C.36.3

3 P.1 and descendent (Gamma), P.1.7 (Gamma+), B.1.1.529 (Omicron), A.27, A.28, B.1.616, B.1.630, C.1.2

4 B.1.351 + L18F (Beta+)

5 B.1.617.2 (Delta), AY lineages (Delta+), AY.34 (Delta+), B.1.617.2 + E484X (Delta+), B.1.617.2 + Q613H (Delta+), B.1.1.529 (Omicron), AY.4.2, B.1.1.519

6 B.1.351 and descendent (Beta), B.1.351 + L18F (Beta+), B.1.351 + P384L (Beta+), B.1.351 + E516Q (Beta+), B.1.526 (Iota)

