## Supplementary Information for "Early detection and improved genomic surveillance of SARS-CoV-2 variants from deep sequencing data"

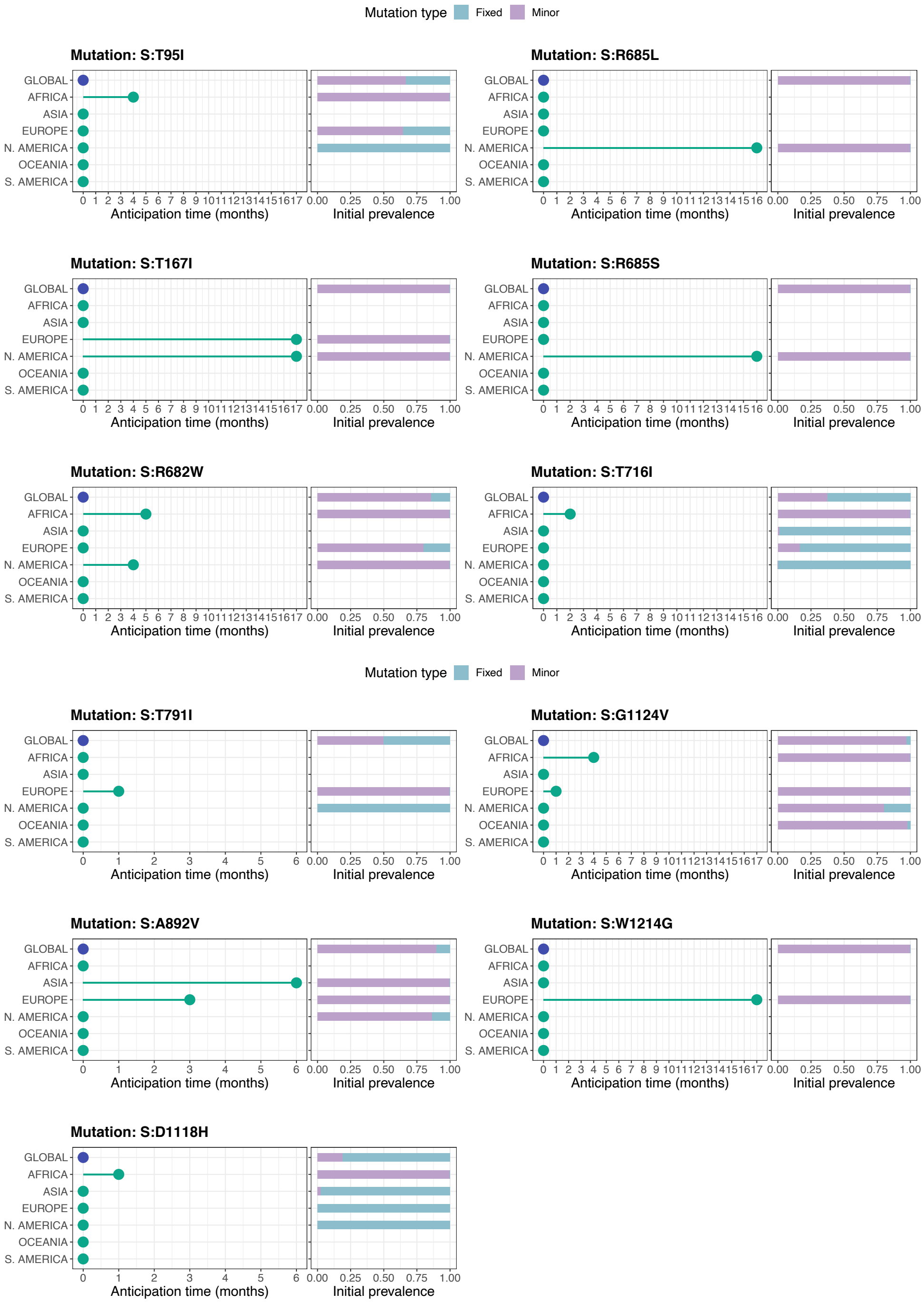


Figure 1: **Early detection of 11 highly diffused S mutations from deep sequencing data.** Analysis of S mutations: S:T95I, S:T167I, S:R682W, S:R685L, S:R685S, S:T716I, S:T791I, S:A892V, S:D1118H, S:G1124V, S:W1214G. The leftmost panels report how many months in advance any mutation was detected as minor (MF $\geq$ 5% and $<$ 50%) in at least 5 samples, while been undetected as fixed (MF $\geq$ 50%), either at the global scale or in any of the 6 considered geographical regions. The rightmost panels show the proportion of samples exhibiting the mutation either as minor or fixed, in the first month in which the mutation was firstly detected in at least 5 samples, either at the global or the local scale.

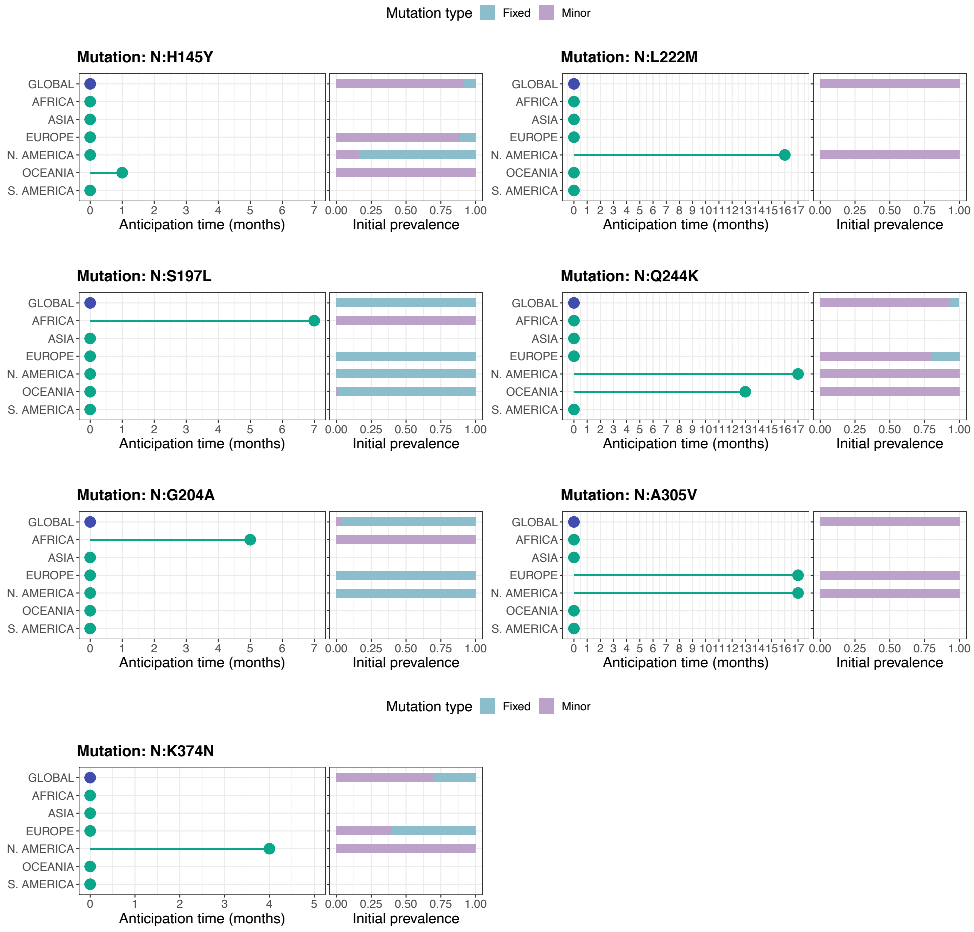


Figure 2: **Early detection of 7 highly diffused N mutations from deep sequencing data.** Analysis of N mutations: N:H145Y, N:S197L, N:G204A, N:L222M, N:Q244K, N:A305V, N:K374N. The leftmost panels report how many months in advance any mutation was detected as minor (MF $\geq 5\%$ and $< 50\%$) in at least 5 samples, while been undetected as fixed (MF $\geq 50\%$), either at the global scale or in any of the 6 considered geographical regions. The rightmost panels show the proportion of samples exhibiting the mutation either as minor or fixed, in the first month in which the mutation was firstly detected in at least 5 samples, either at the global or the local scale.

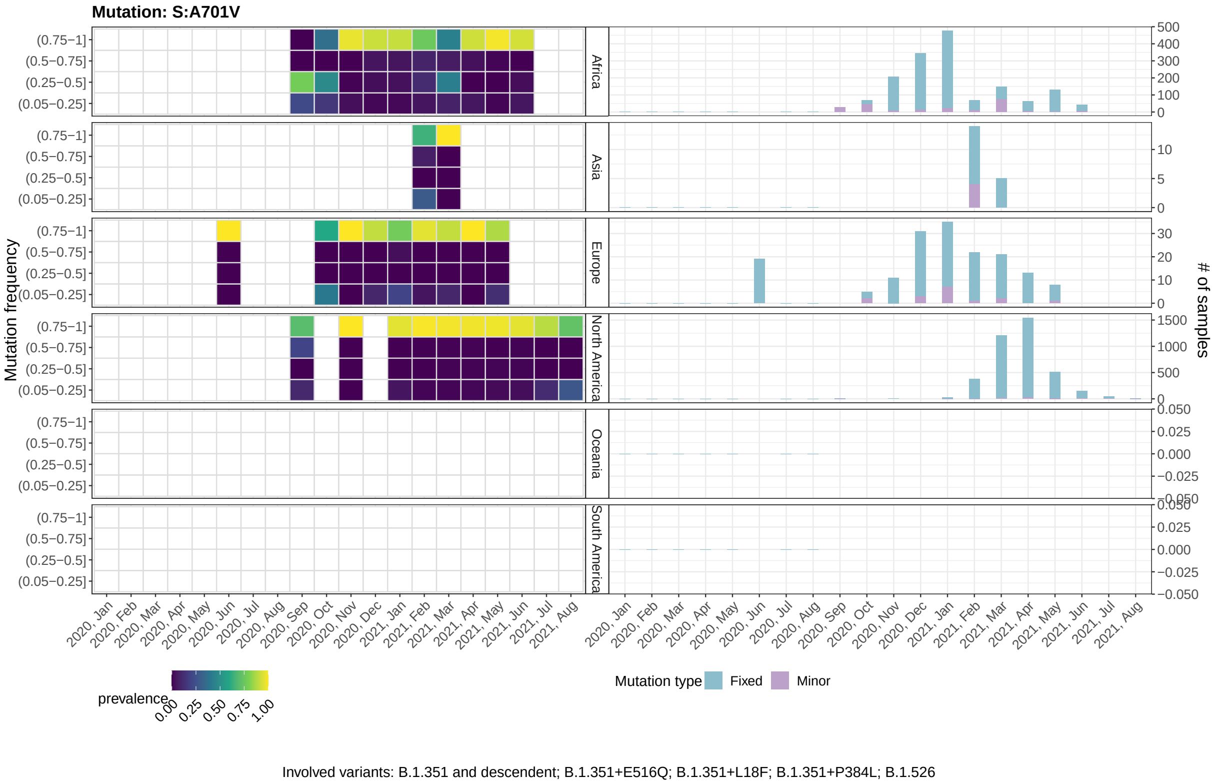


Figure 3: **Mutant frequency and prevalence variation in time of mutation: S:A701V.** The leftmost panels return the distribution of the mutation frequency (MF) of all samples with the mutation, grouped by month and geographical region. Each cell shows the proportion of samples showing the mutation with that specific MF. The rightmost panels show the number of samples showing the mutations either as minor (MF $\geq 5\%$ and $< 50\%$) or fixed (MF $\geq 50\%$).

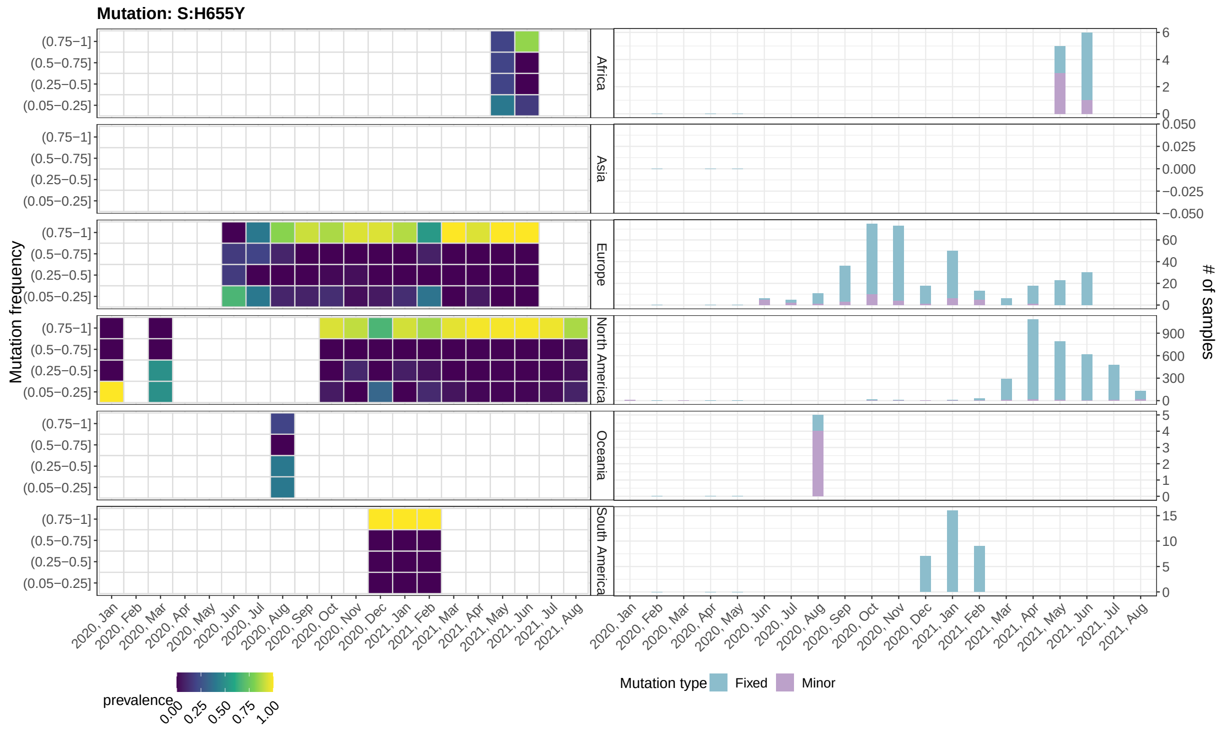


Involved variants: A.27; A.28; B.1.616; B.1.630; C.1.2; P.1 and descendent; P.1.7

Figure 4: **Mutant frequency and prevalence variation in time of mutation: S:H655Y.** Please refer to the caption of Supplementary Figure 3

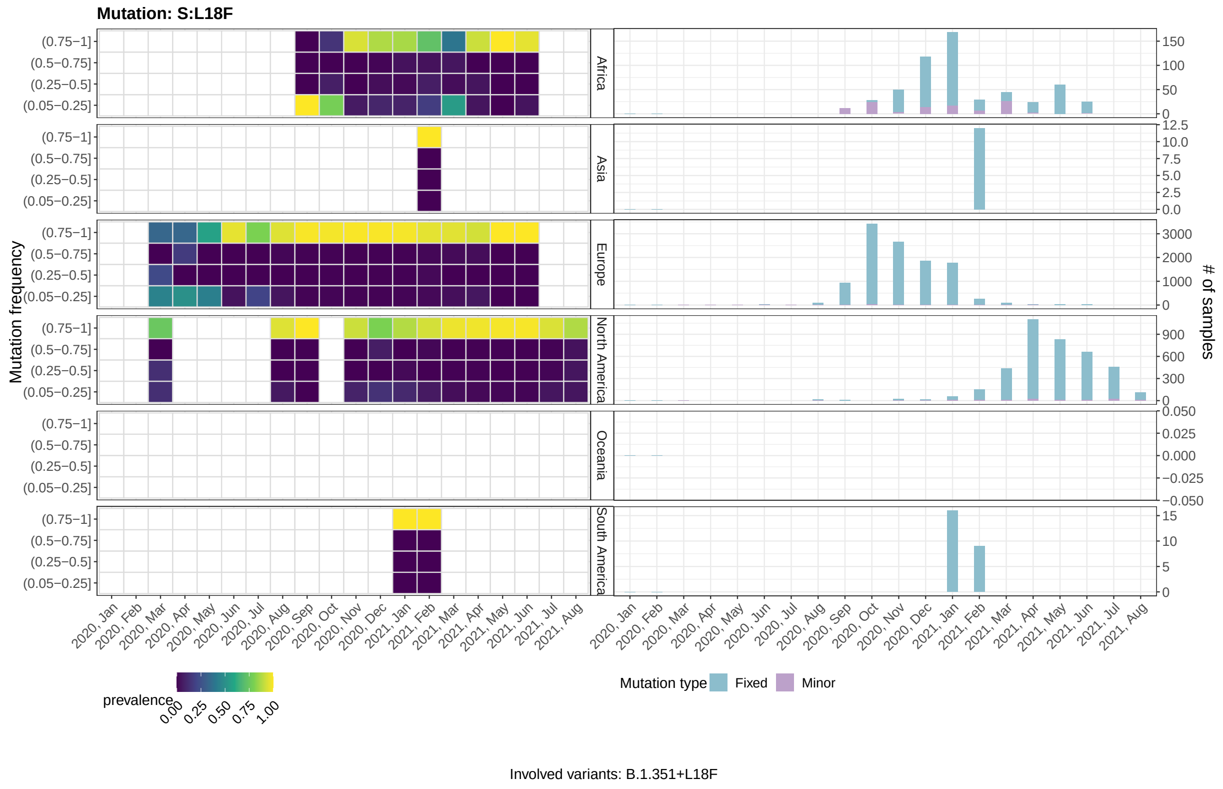


Figure 5: **Mutant frequency and prevalence variation in time of mutation: S:L18F.** Please refer to the caption of Supplementary Figure 3

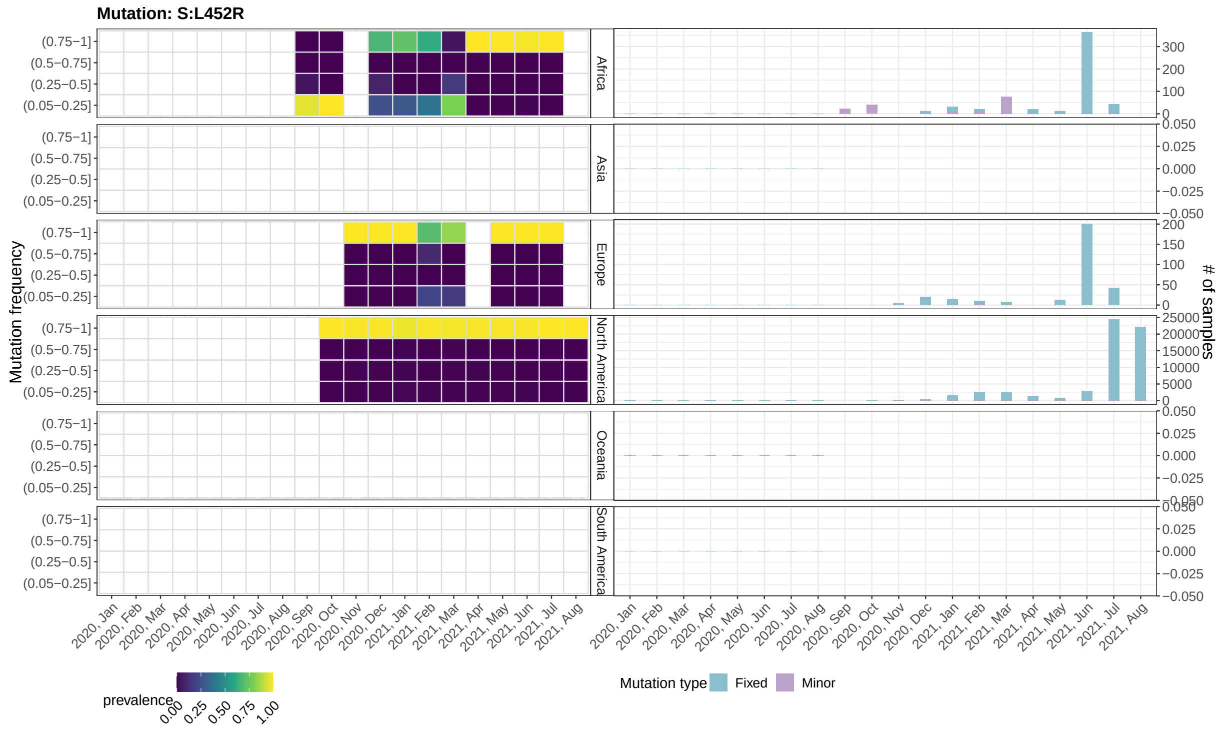


Involved variants: A.27; AY.34; AY.4.2; B.1.1.7+L452R; B.1.427|B.1.429|C.16; B.1.617.1|B.1.617.3; B.1.617.2; B.1.617.2 descendent; B.1.617.2+E484X; B.1.617.2+Q613H; B.1.630 C.36.3

Figure 6: **Mutant frequency and prevalence variation in time of mutation: S:L452R.** Please refer to the caption of Supplementary Figure 3

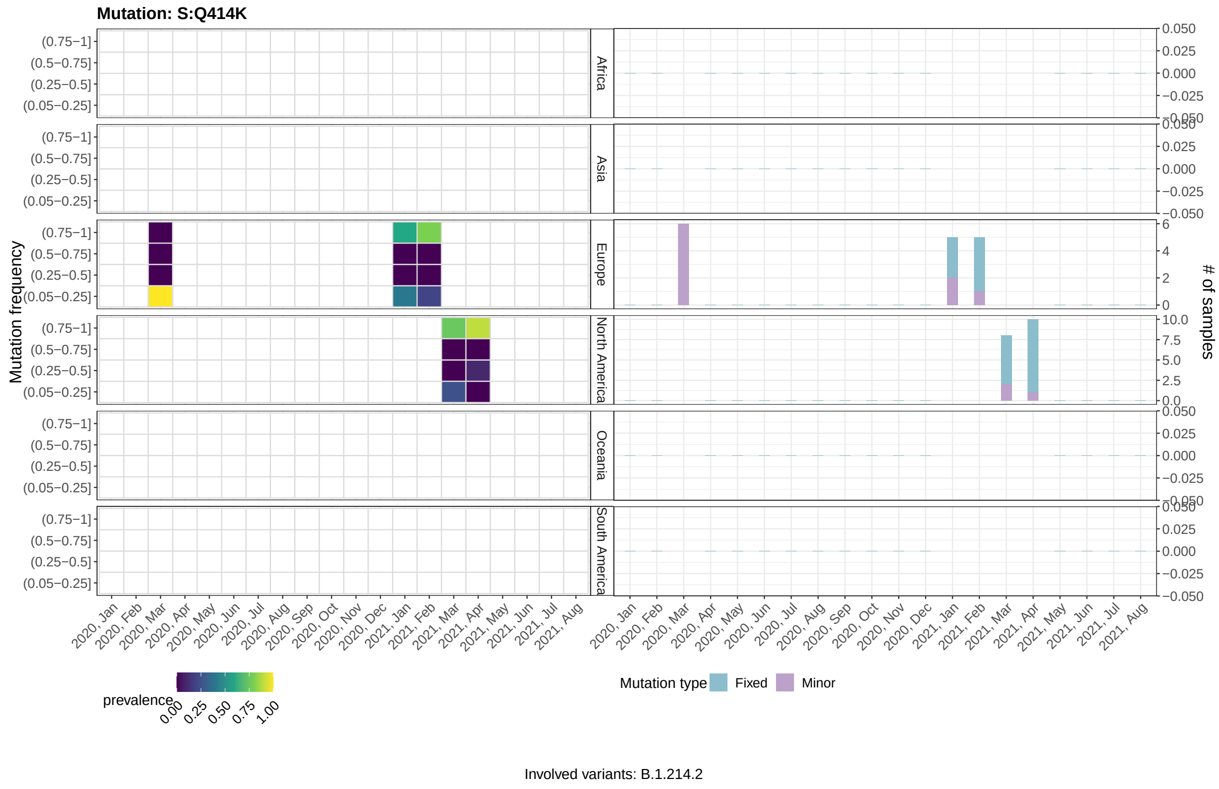


Figure 7: **Mutant frequency and prevalence variation in time of mutation: S:Q414K.** Please refer to the caption of Supplementary Figure 3

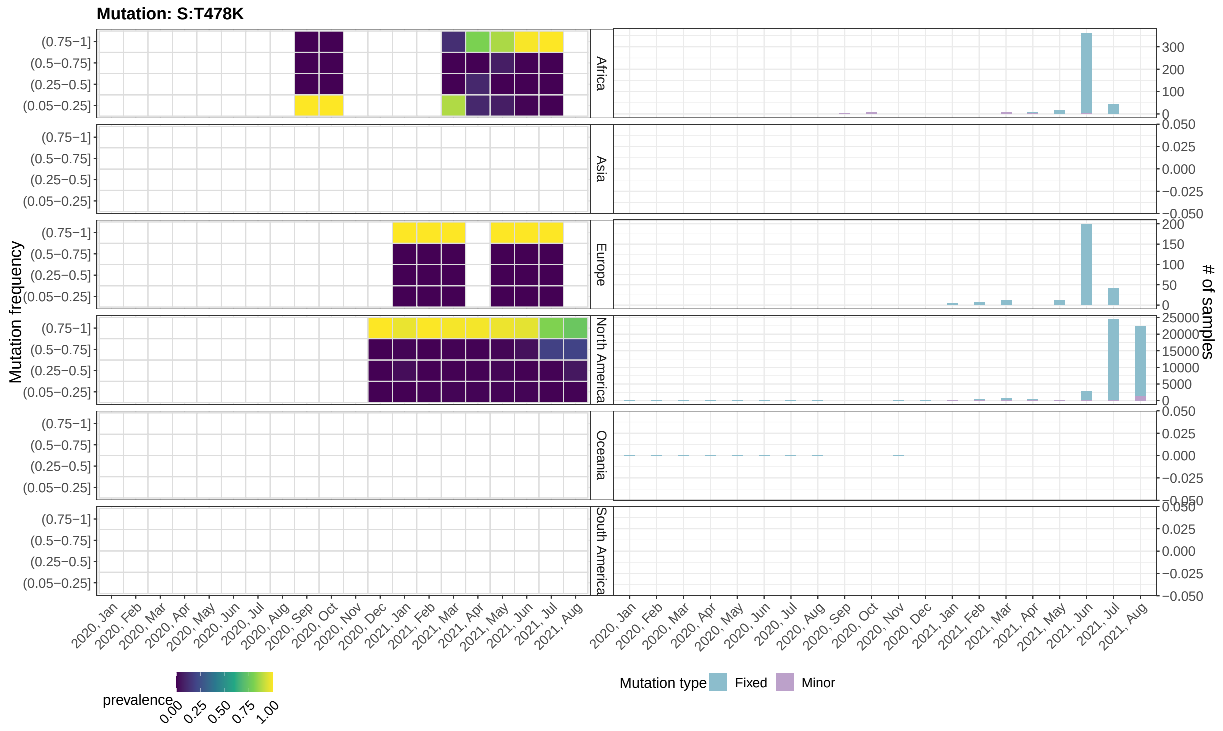


Involved variants: AY.34; AY.4.2; B.1.1.519; B.1.617.2; B.1.617.2 descendent; B.1.617.2+E484X; B.1.617.2+Q613H

Figure 8: **Mutant frequency and prevalence variation in time of mutation: S:T478K.** Please refer to the caption of Supplementary Figure 3

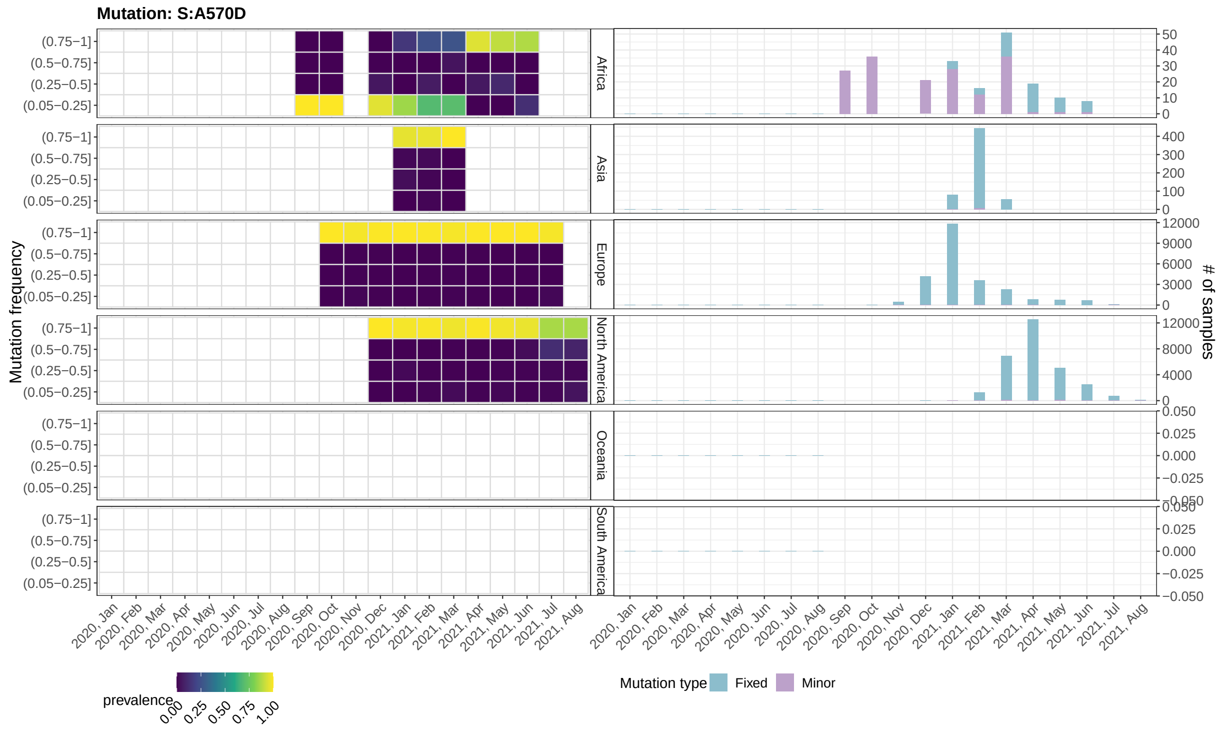


Figure 9: **Mutant frequency and prevalence variation in time of mutation: S:A570D.** Please refer to the caption of Supplementary Figure 3

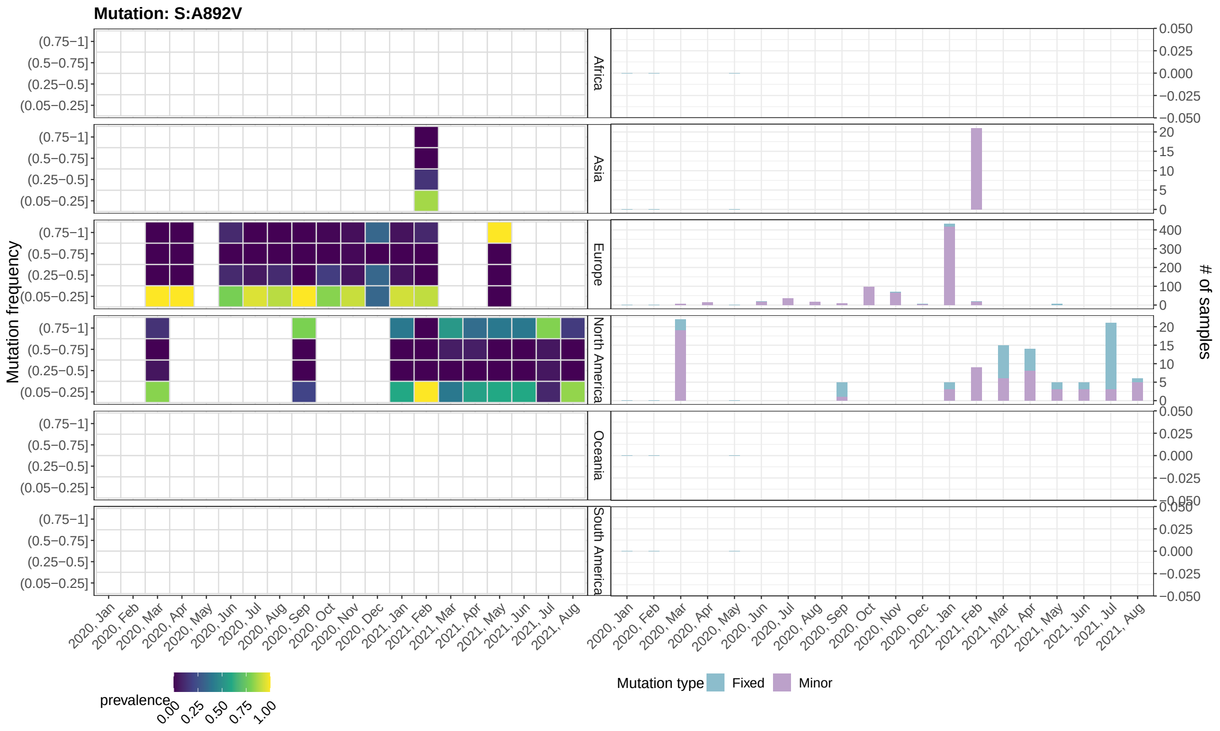


Figure 10: **Mutant frequency and prevalence variation in time of mutation: S:A892V.** Please refer to the caption of Supplementary Figure 3

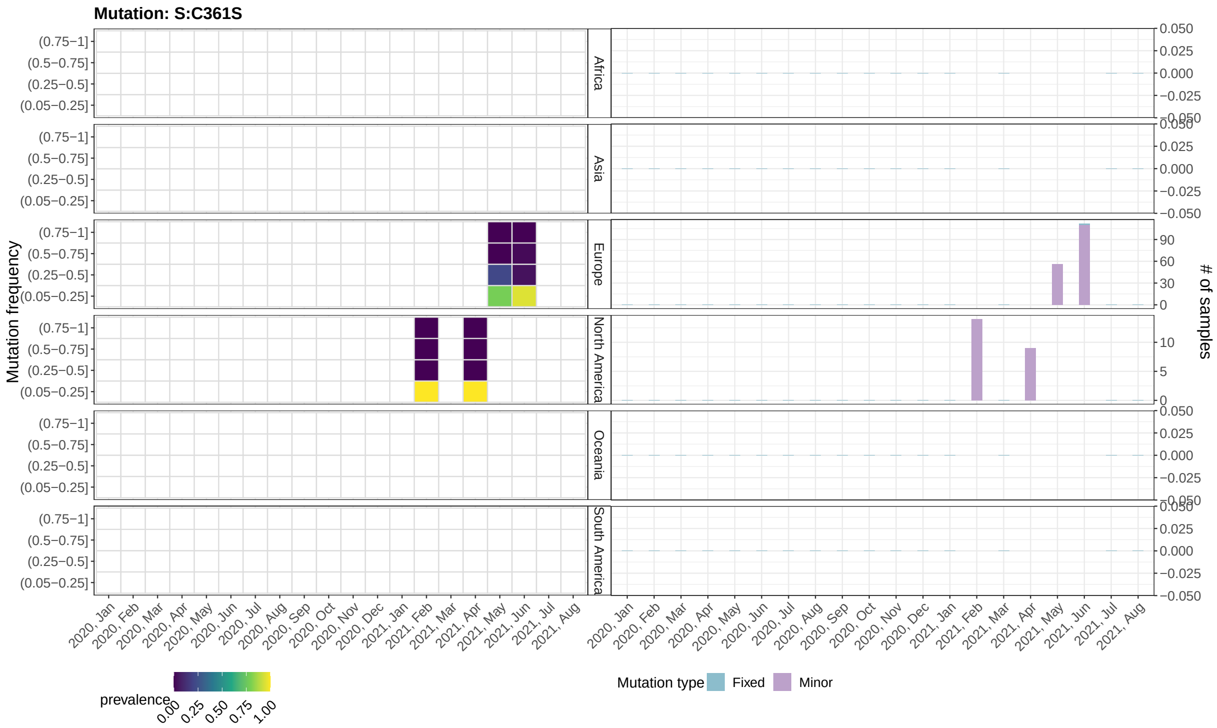


Figure 11: **Mutant frequency and prevalence variation in time of mutation: S:C361S.** Please refer to the caption of Supplementary Figure 3

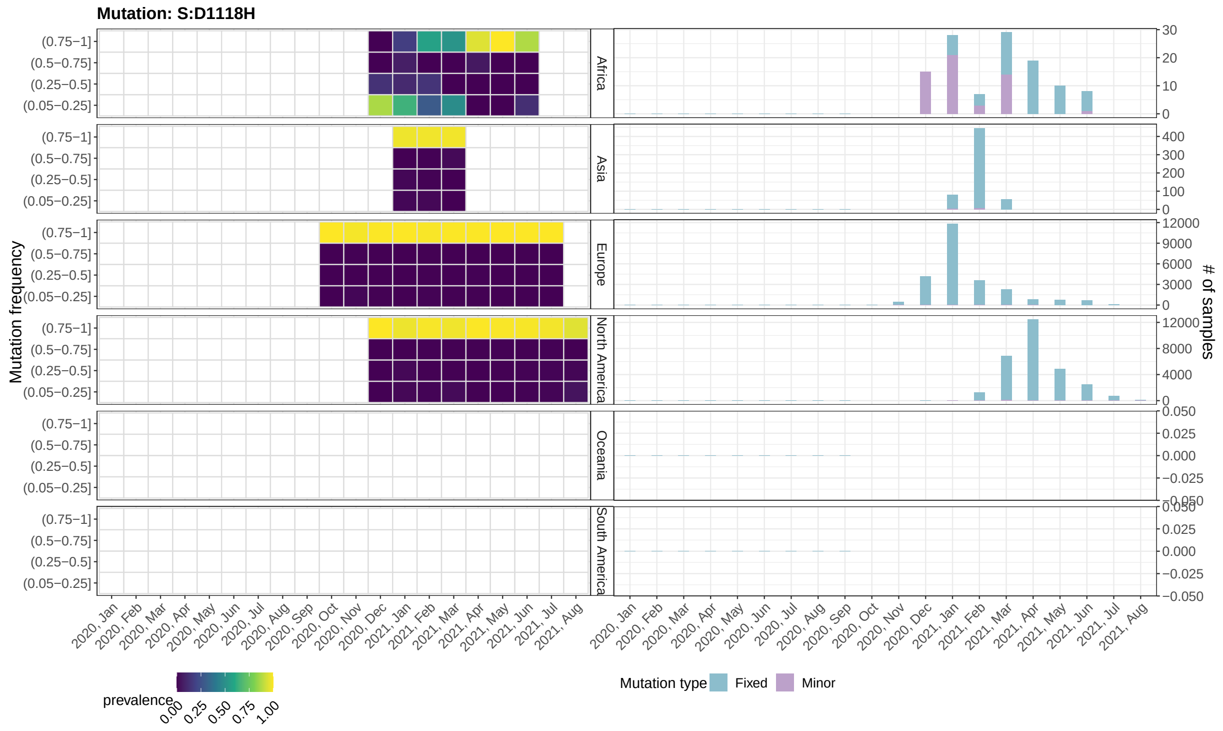


Figure 12: **Mutant frequency and prevalence variation in time of mutation: S:D1118H.** Please refer to the caption of Supplementary Figure 3

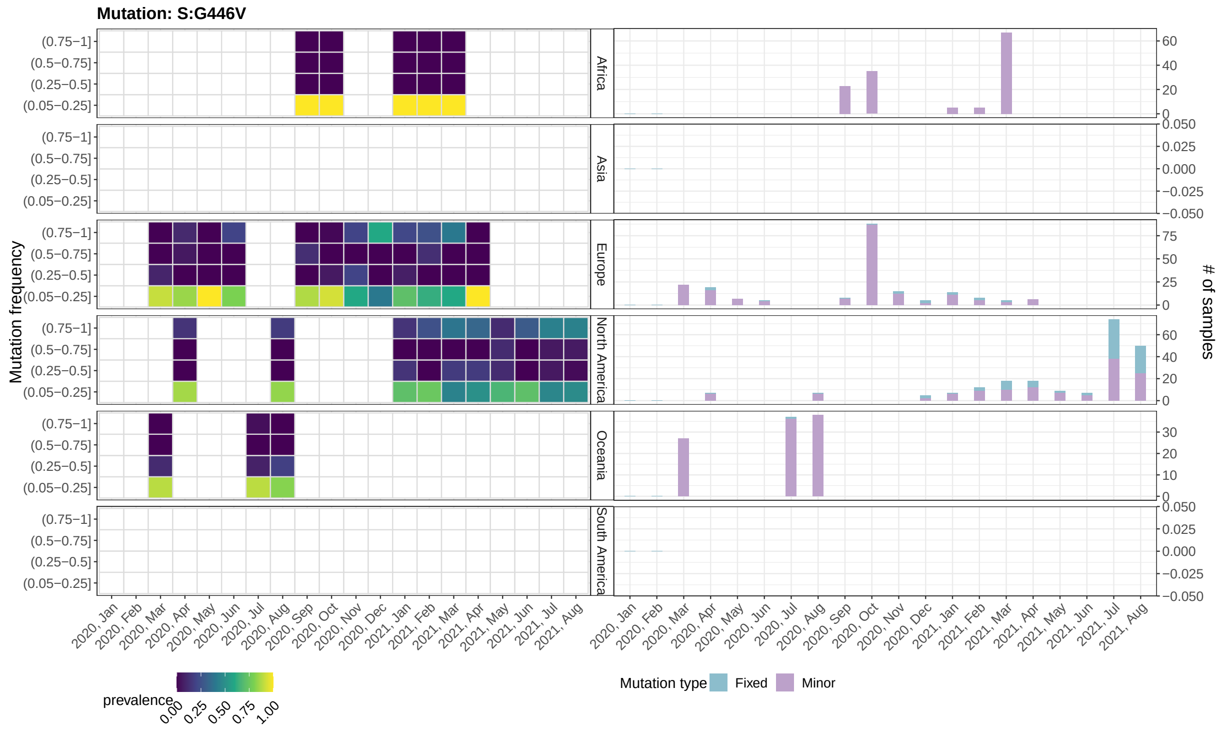


Figure 13: **Mutant frequency and prevalence variation in time of mutation: S:G446V.** Please refer to the caption of Supplementary Figure 3

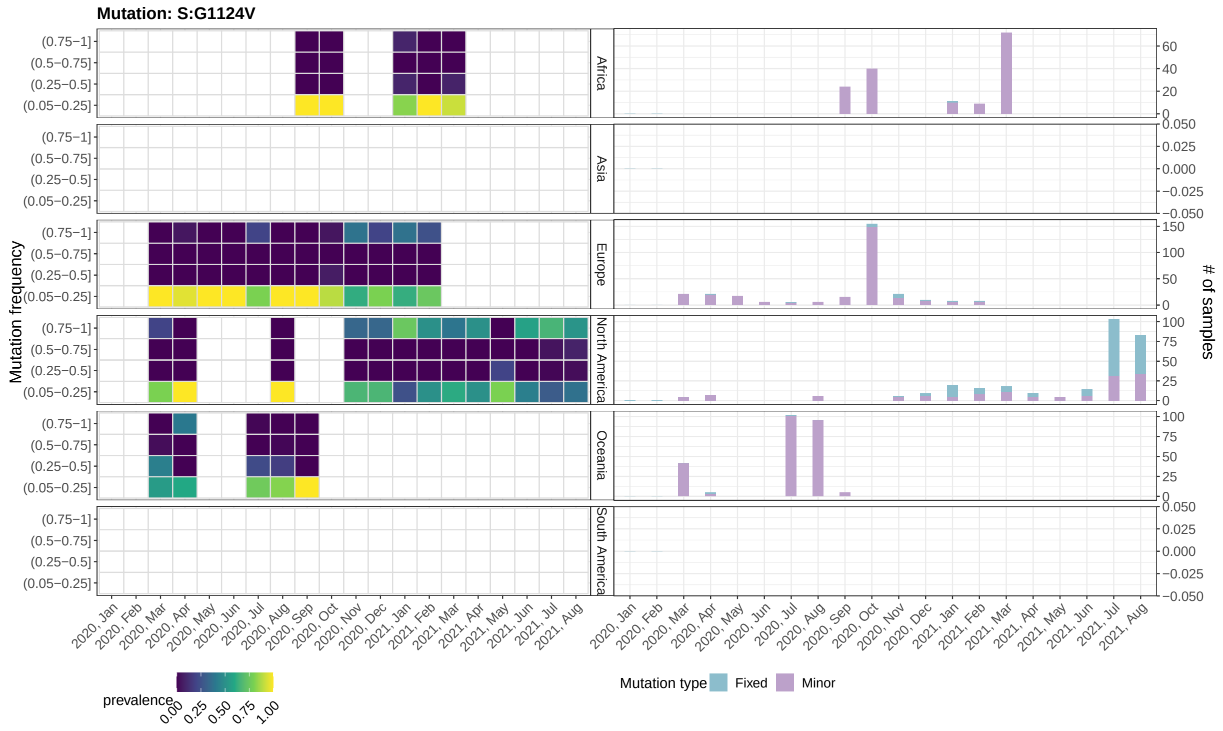


Figure 14: **Mutant frequency and prevalence variation in time of mutation: S:G1124V.** Please refer to the caption of Supplementary Figure 3

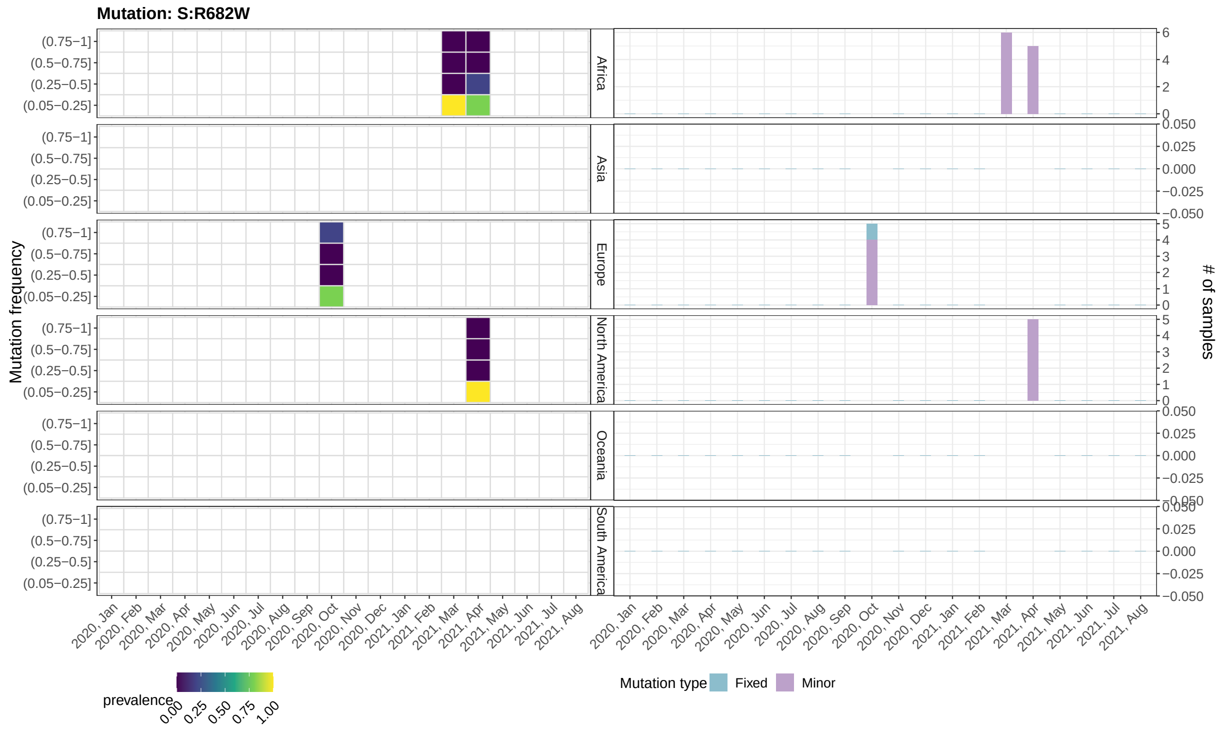


Figure 15: **Mutant frequency and prevalence variation in time of mutation: S:R682W.** Please refer to the caption of Supplementary Figure 3

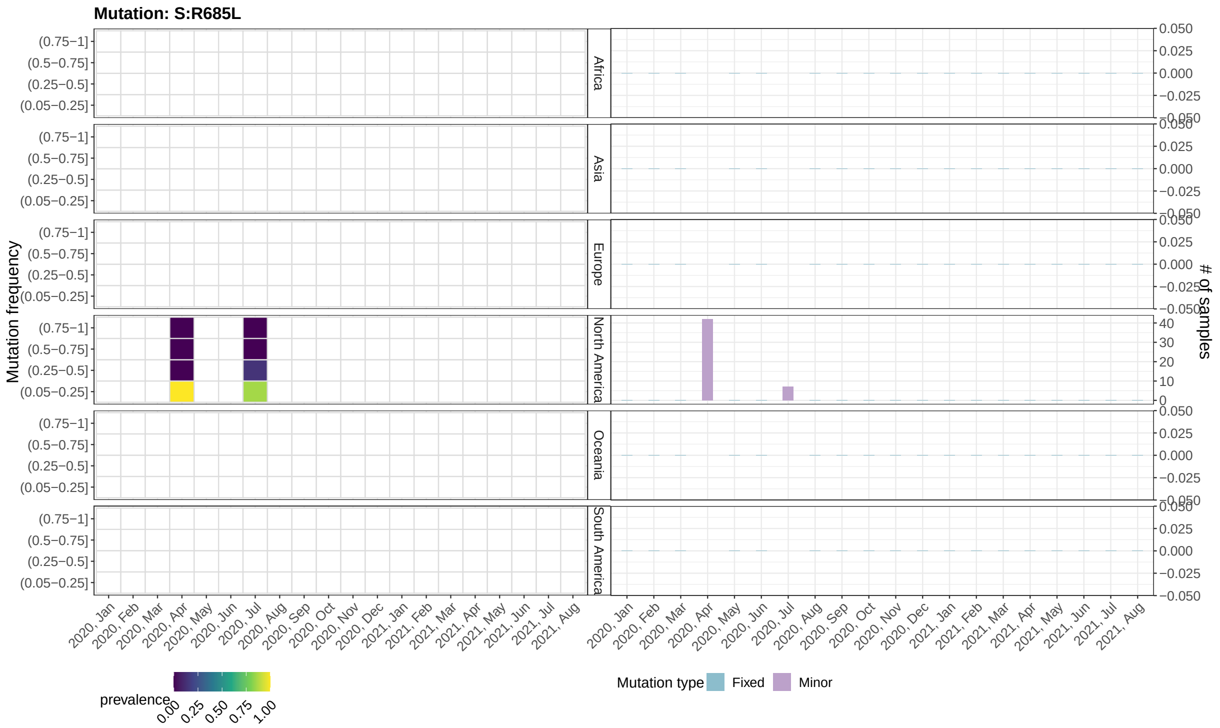


Figure 16: **Mutant frequency and prevalence variation in time of mutation: S:R685L.** Please refer to the caption of Supplementary Figure 3

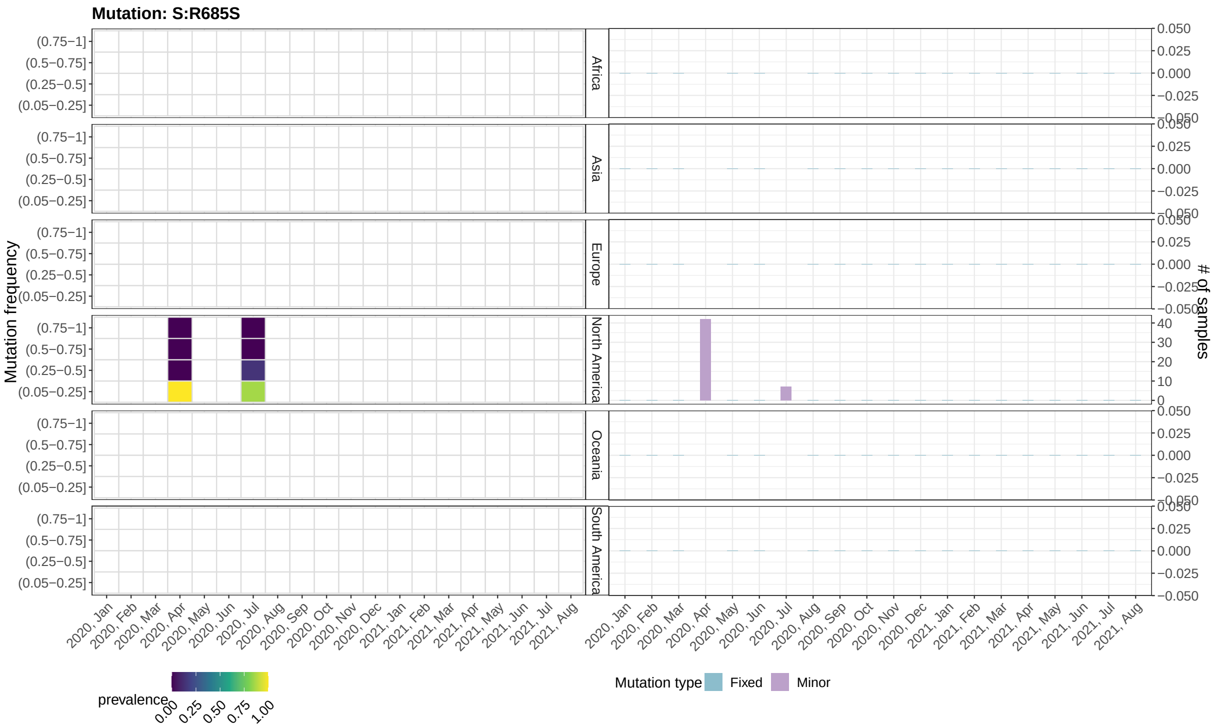


Figure 17: **Mutant frequency and prevalence variation in time of mutation: S:R685S.** Please refer to the caption of Supplementary Figure 3

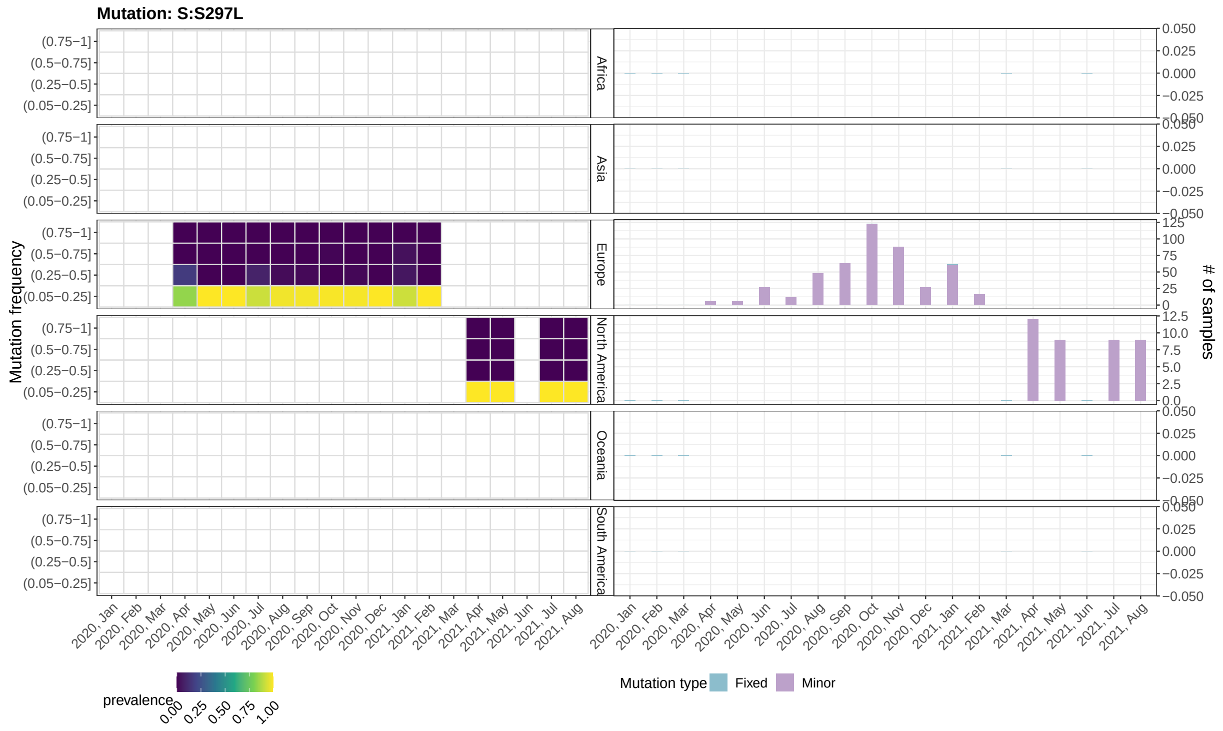


Figure 18: **Mutant frequency and prevalence variation in time of mutation: S:S297L.** Please refer to the caption of Supplementary Figure 3

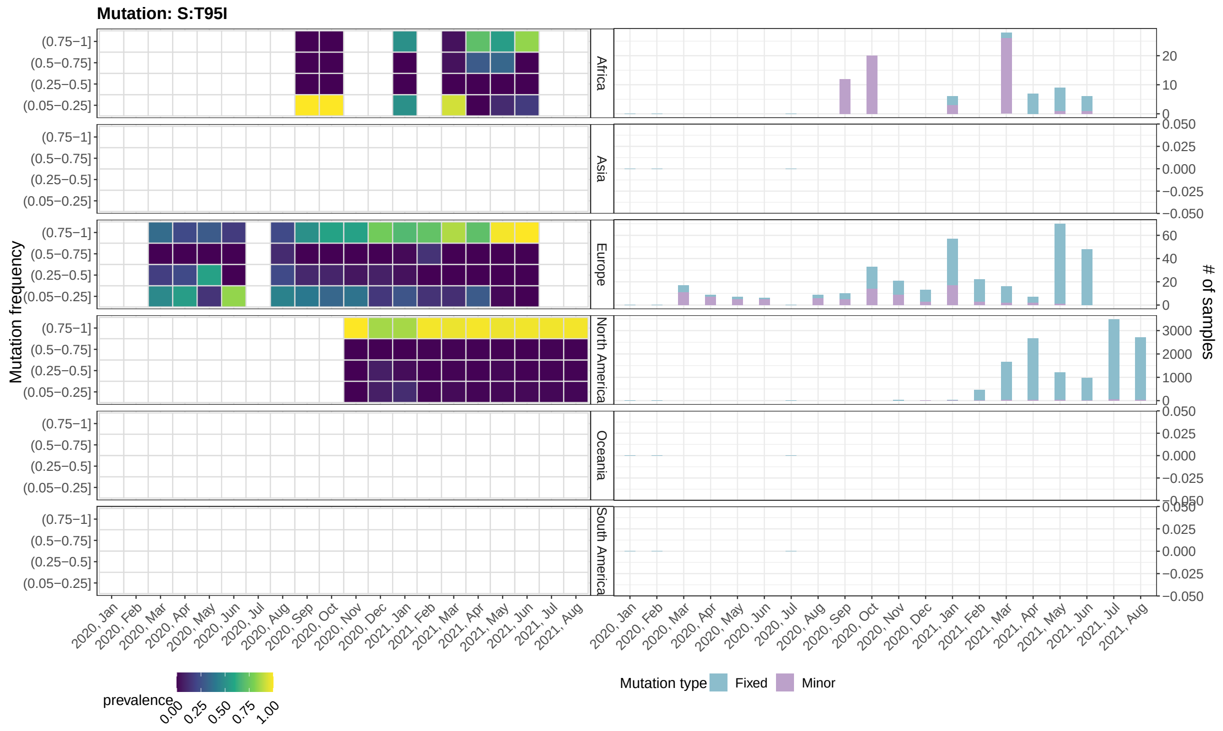


Figure 19: **Mutant frequency and prevalence variation in time of mutation: S:T95I.** Please refer to the caption of Supplementary Figure 3

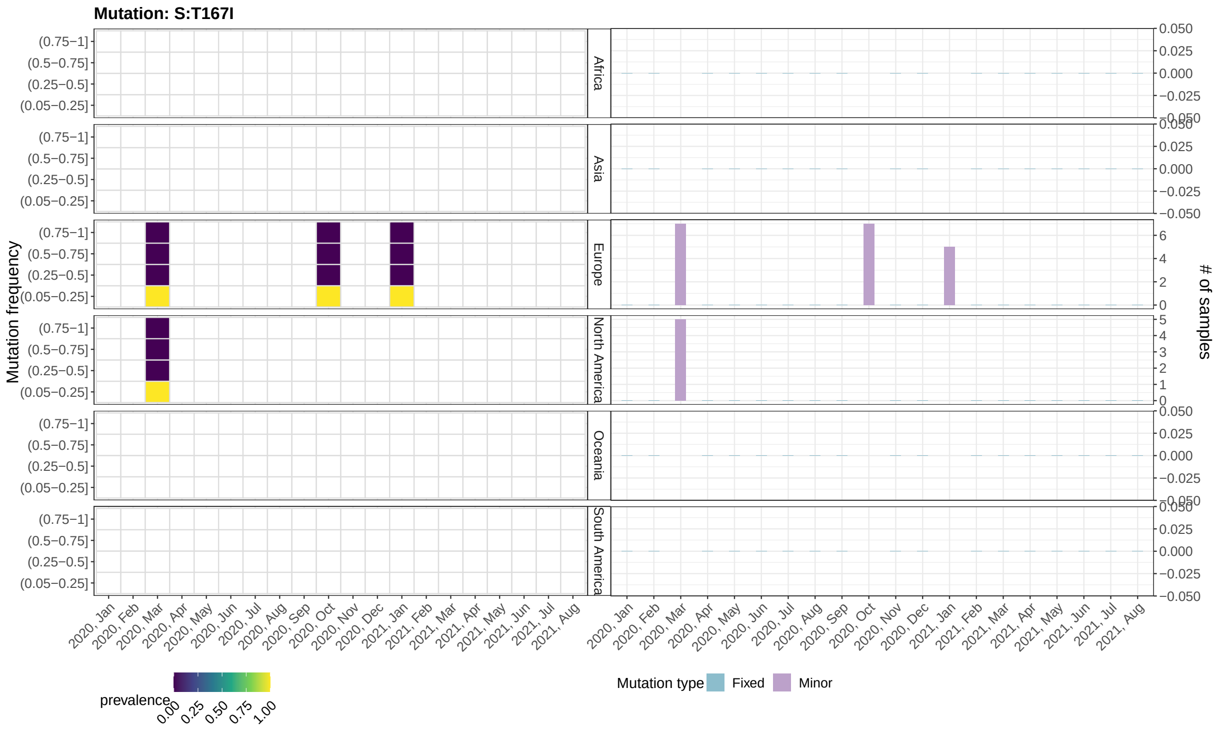


Figure 20: **Mutant frequency and prevalence variation in time of mutation: S:T167I.** Please refer to the caption of Supplementary Figure 3

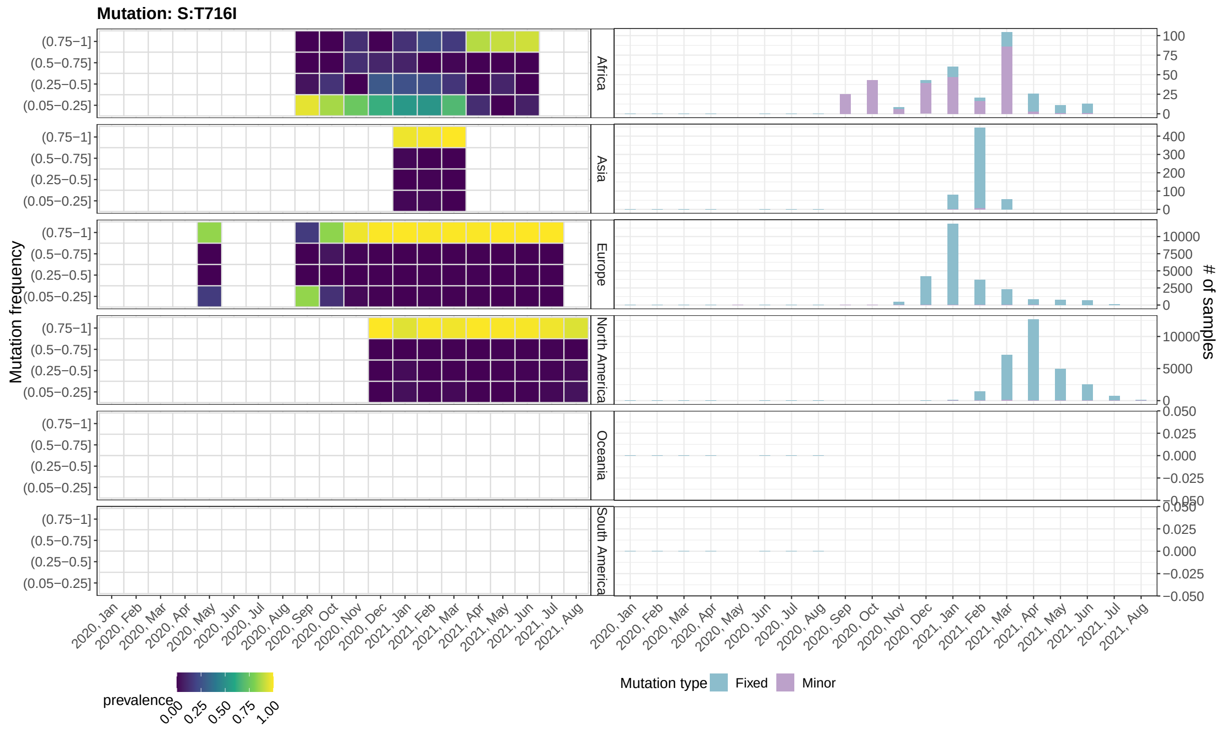


Figure 21: **Mutant frequency and prevalence variation in time of mutation: S:T716I.** Please refer to the caption of Supplementary Figure 3

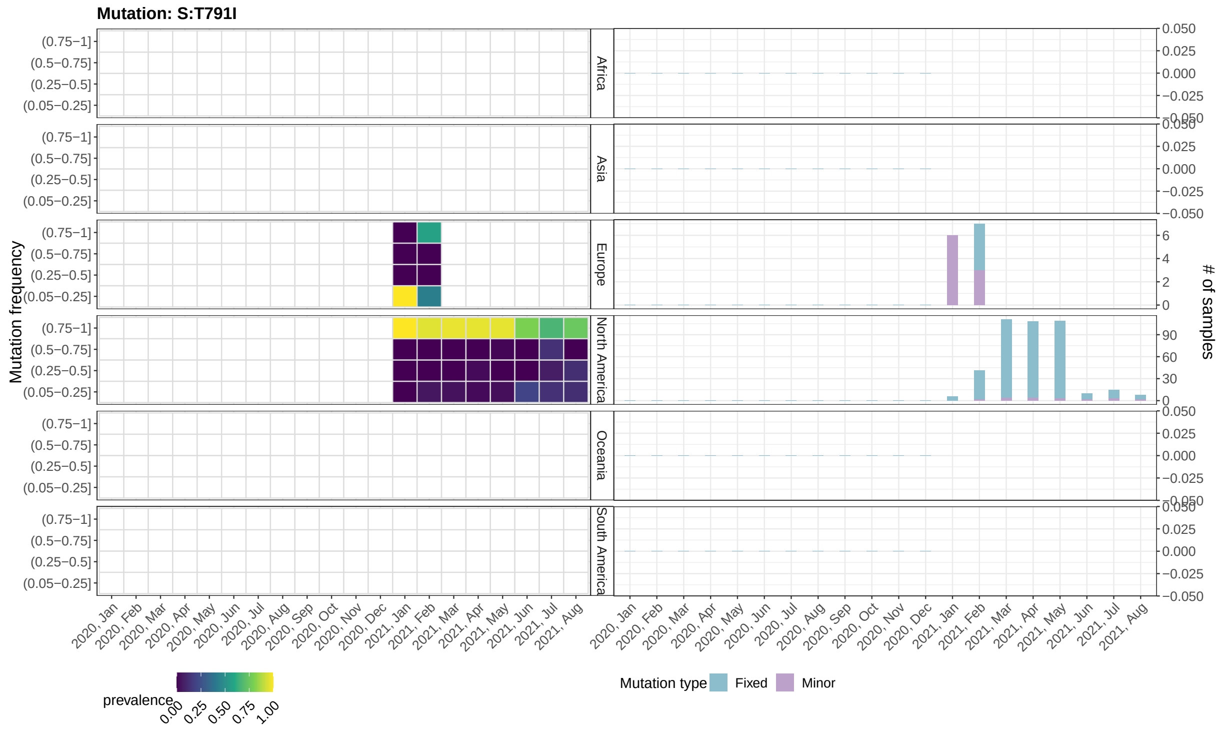


Figure 22: **Mutant frequency and prevalence variation in time of mutation: S:T791I.** Please refer to the caption of Supplementary Figure 3

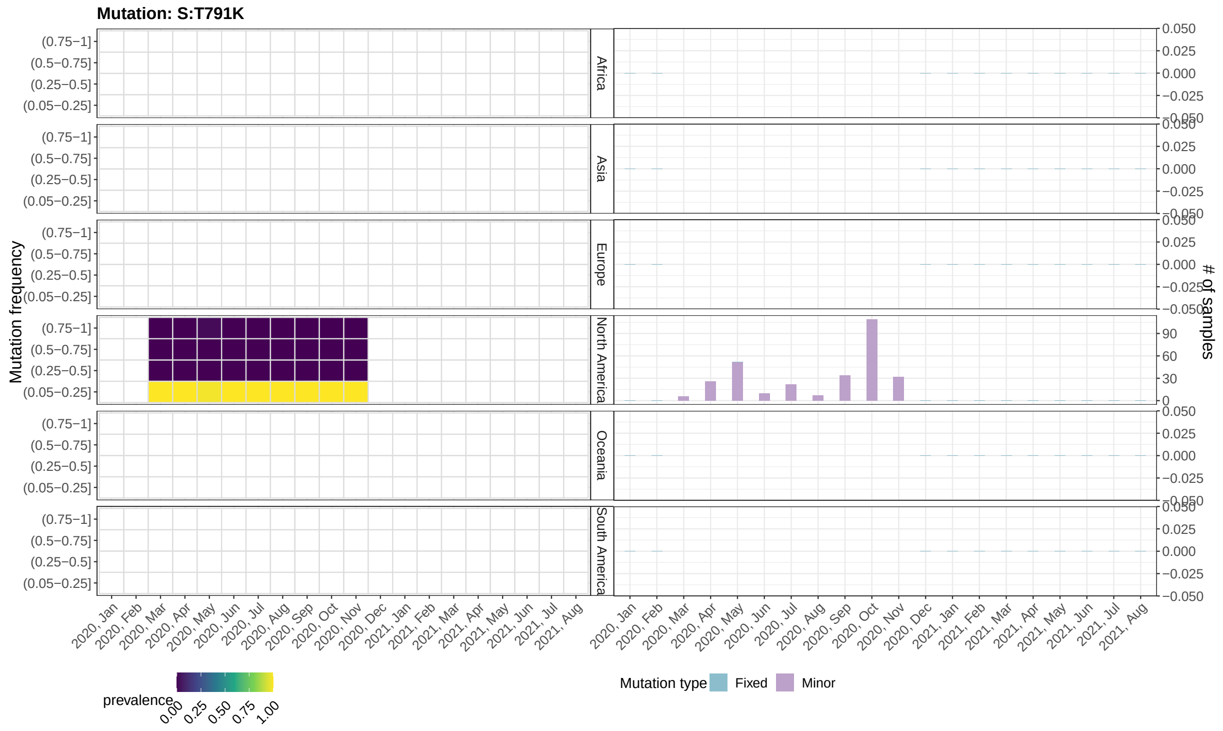


Figure 23: **Mutant frequency and prevalence variation in time of mutation: S:T791K.** Please refer to the caption of Supplementary Figure 3

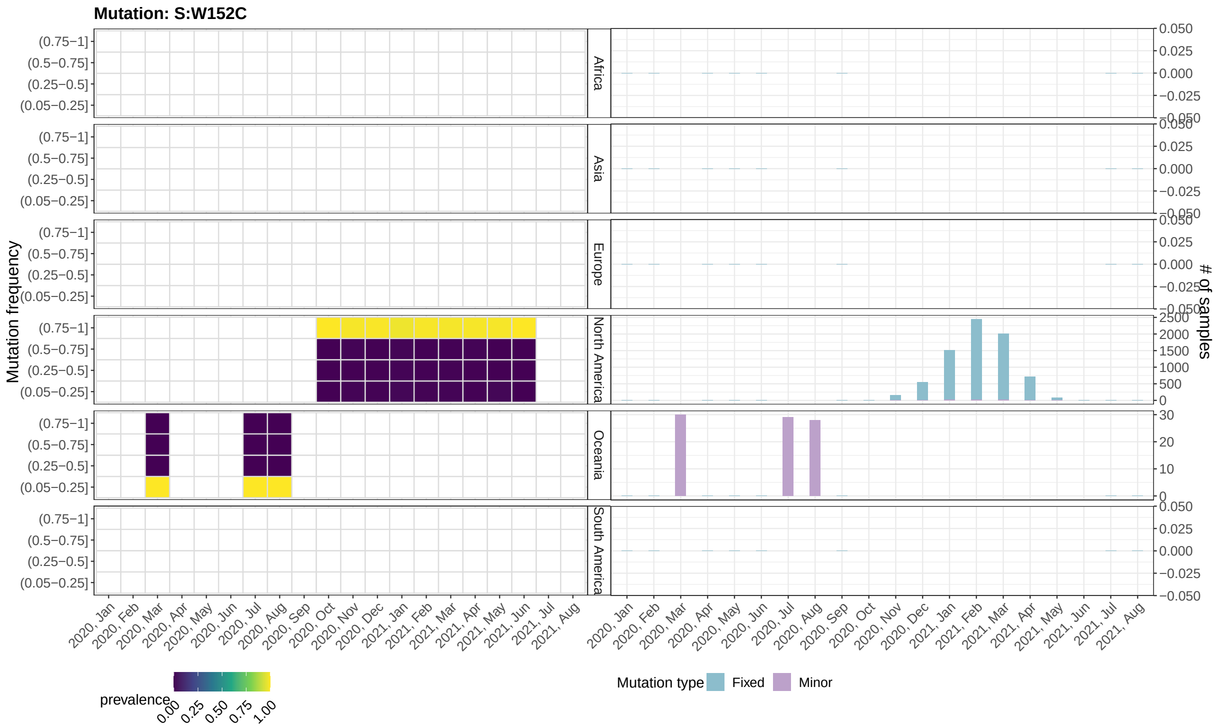


Figure 24: **Mutant frequency and prevalence variation in time of mutation: S:W152C.** Please refer to the caption of Supplementary Figure 3

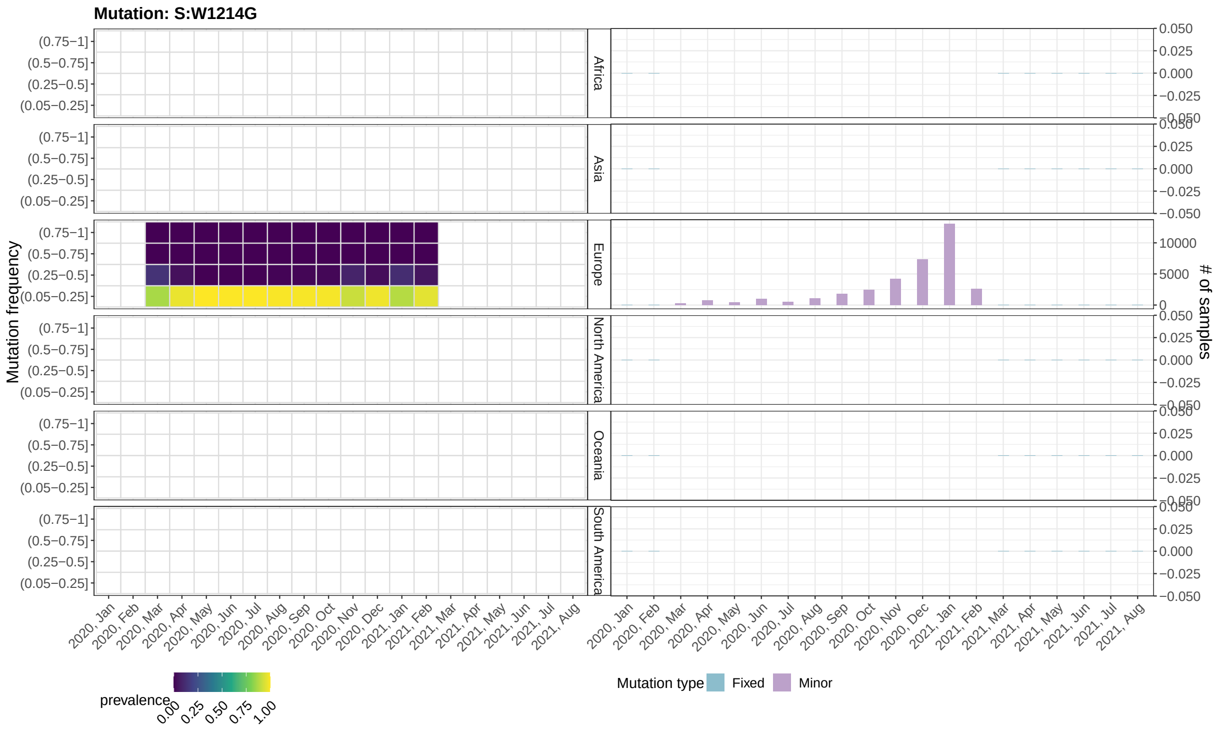


Figure 25: **Mutant frequency and prevalence variation in time of mutation: S:W1214G.** Please refer to the caption of Supplementary Figure 3

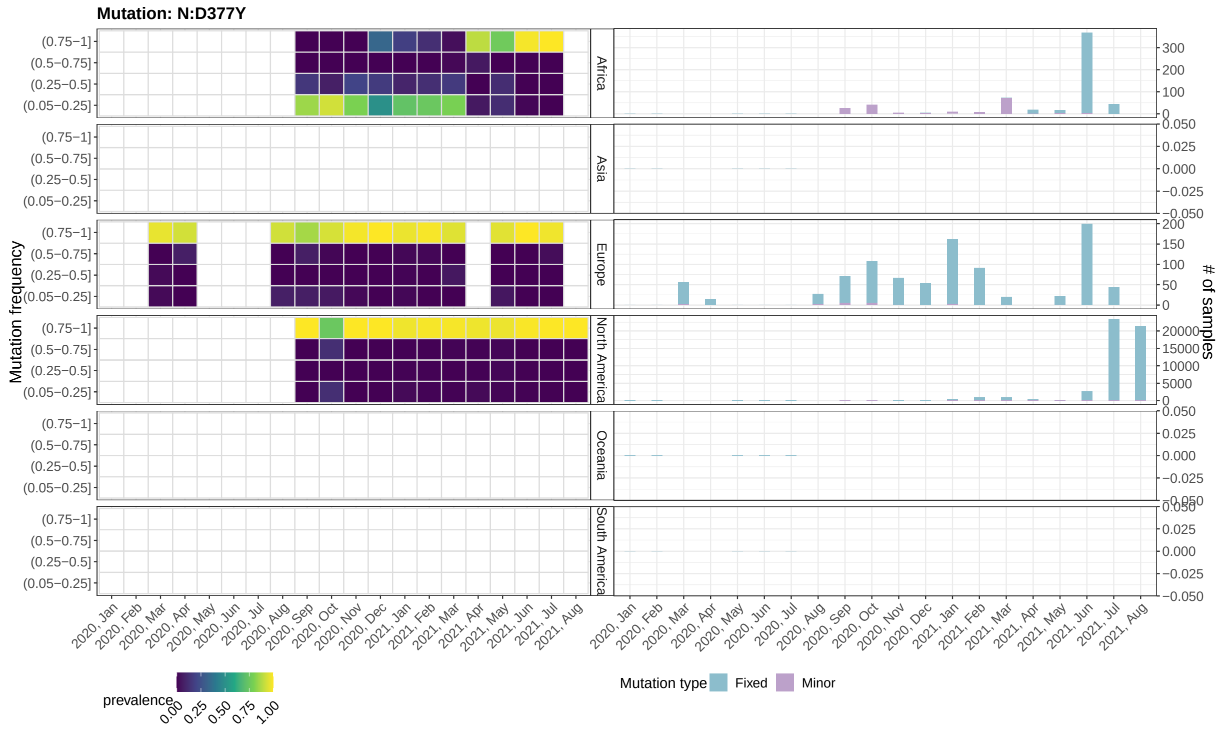


Figure 26: **Mutant frequency and prevalence variation in time of mutation: N:D377Y.** Please refer to the caption of Supplementary Figure 3

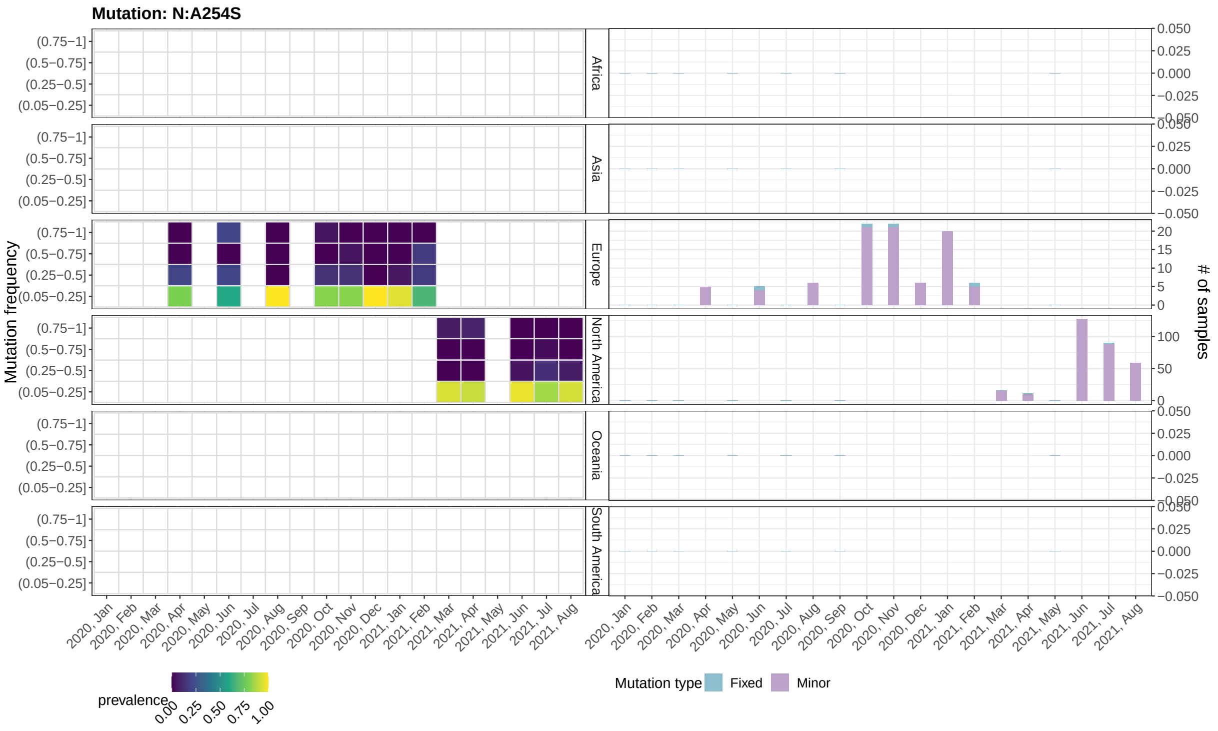


Figure 27: **Mutant frequency and prevalence variation in time of mutation: N:A254S.** Please refer to the caption of Supplementary Figure 3

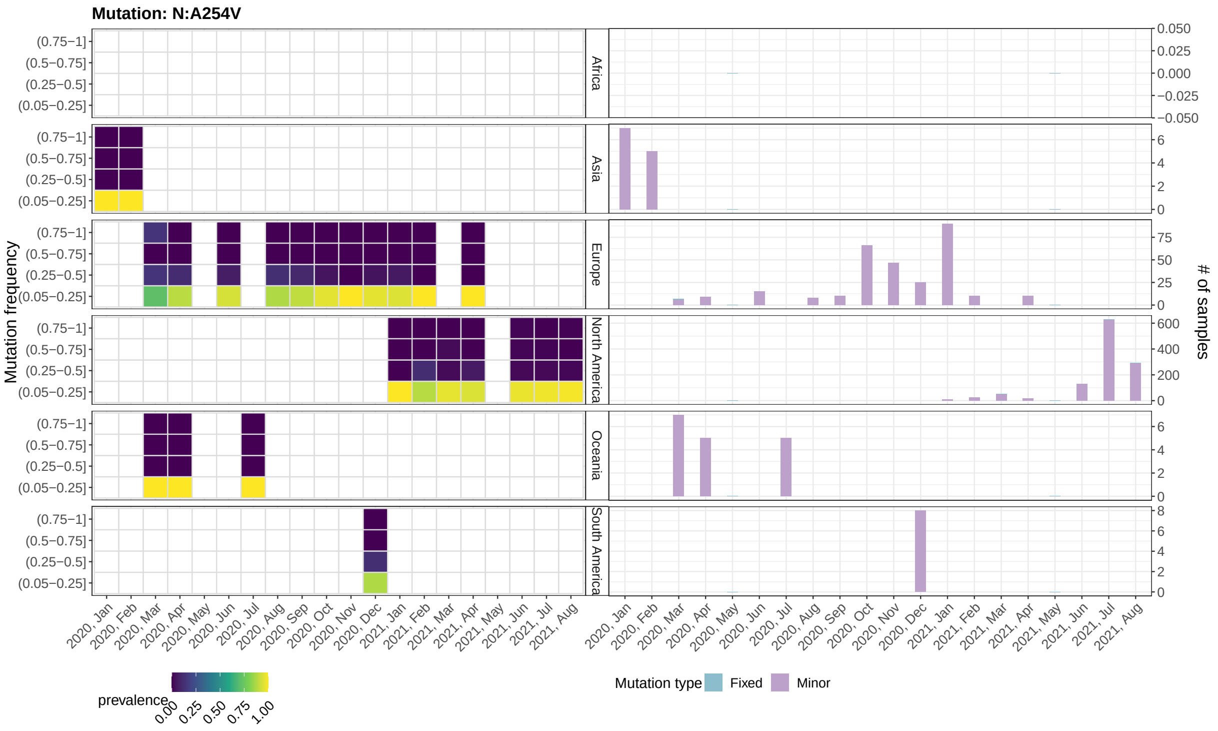


Figure 28: **Mutant frequency and prevalence variation in time of mutation: N:A254V.** Please refer to the caption of Supplementary Figure 3

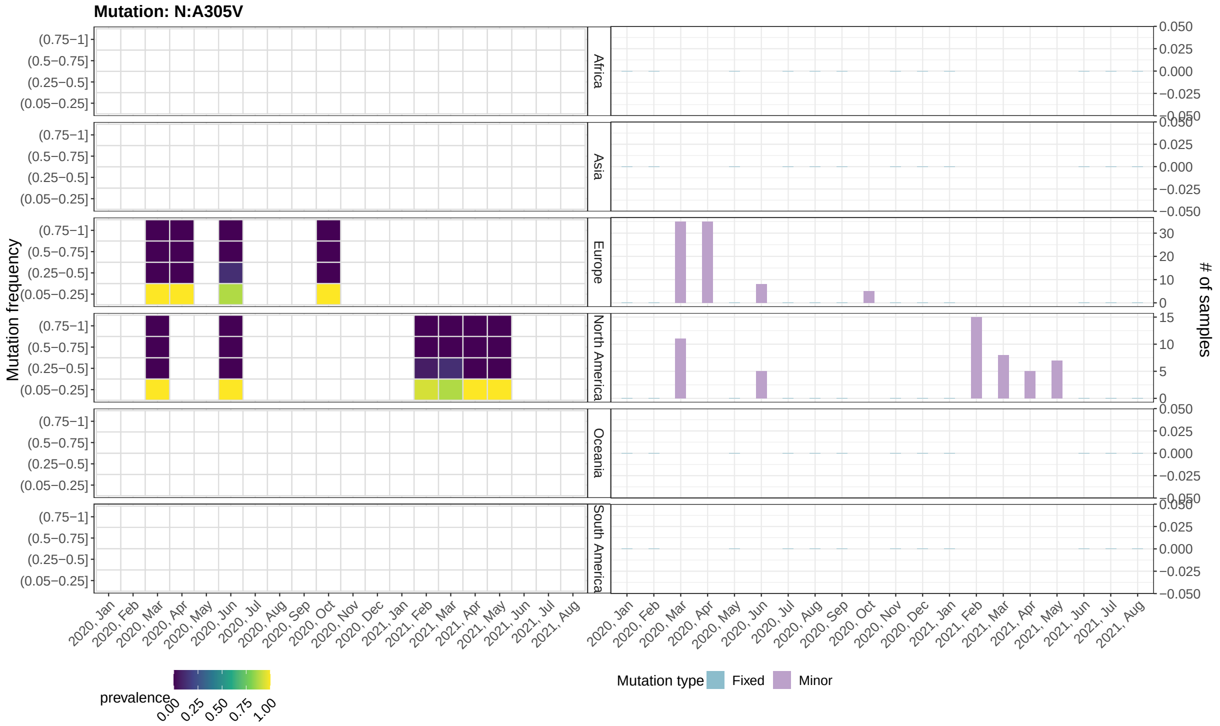


Figure 29: **Mutant frequency and prevalence variation in time of mutation: N:A305V.** Please refer to the caption of Supplementary Figure 3

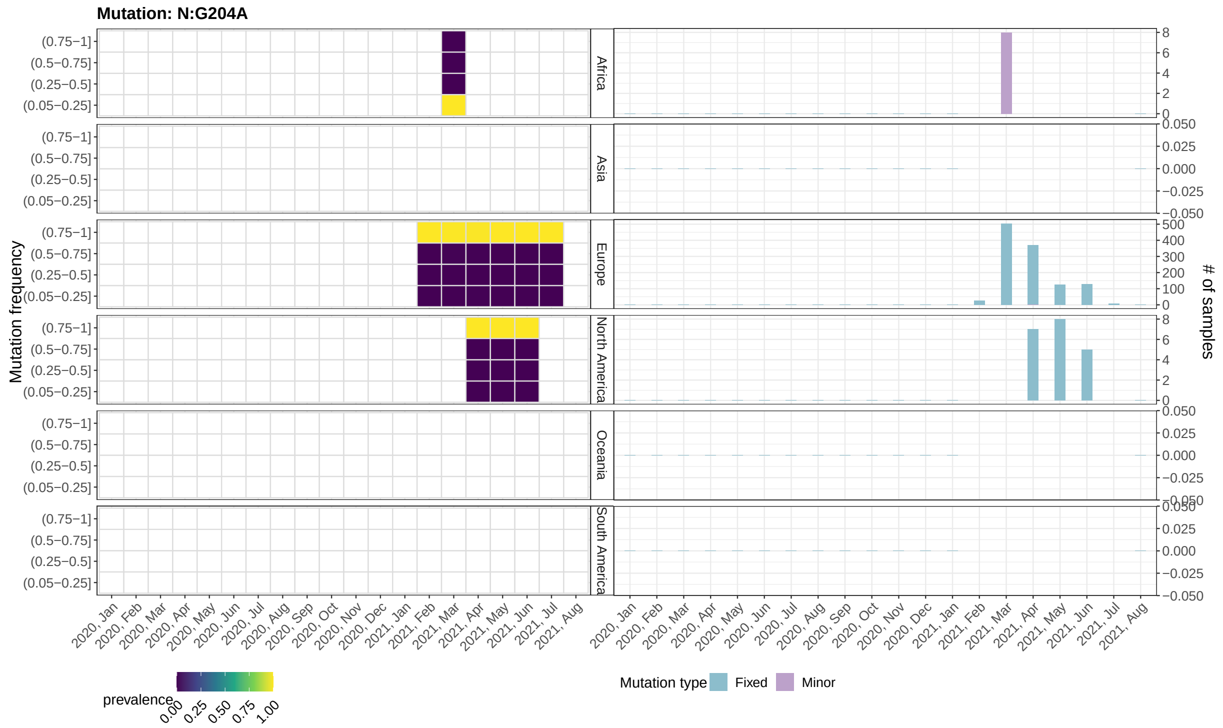


Figure 30: **Mutant frequency and prevalence variation in time of mutation: N:G204A.** Please refer to the caption of Supplementary Figure 3

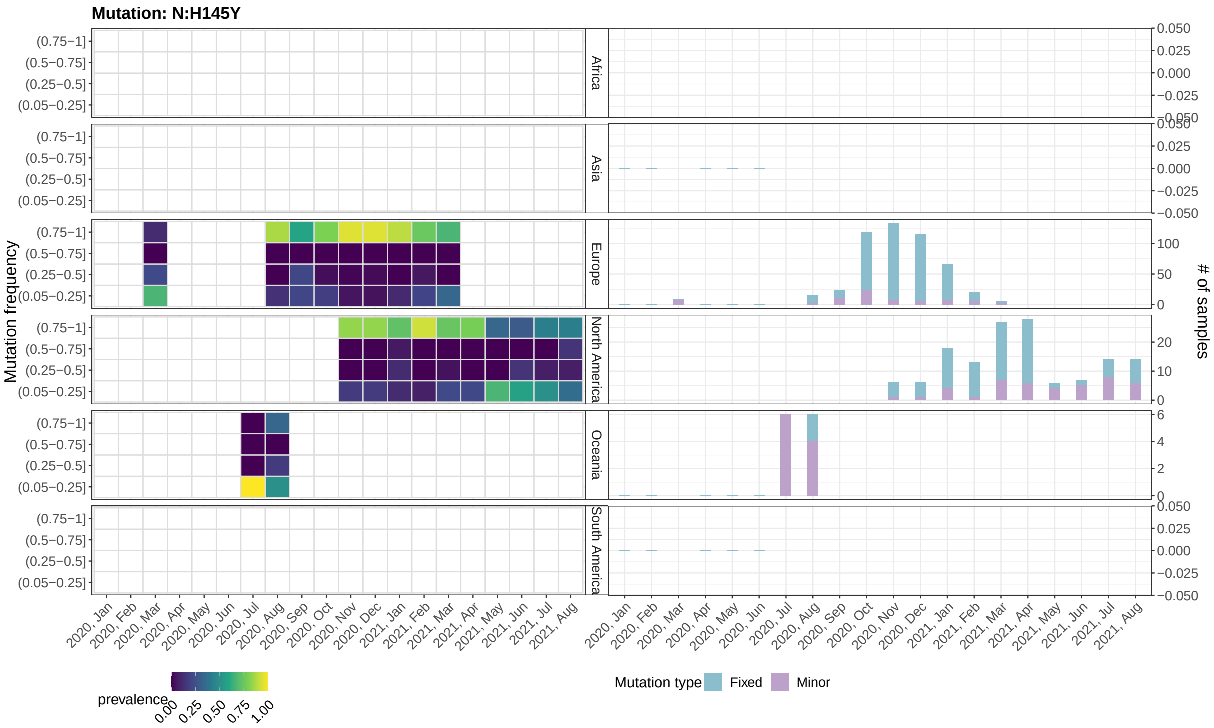


Figure 31: **Mutant frequency and prevalence variation in time of mutation: N:H145Y.** Please refer to the caption of Supplementary Figure 3

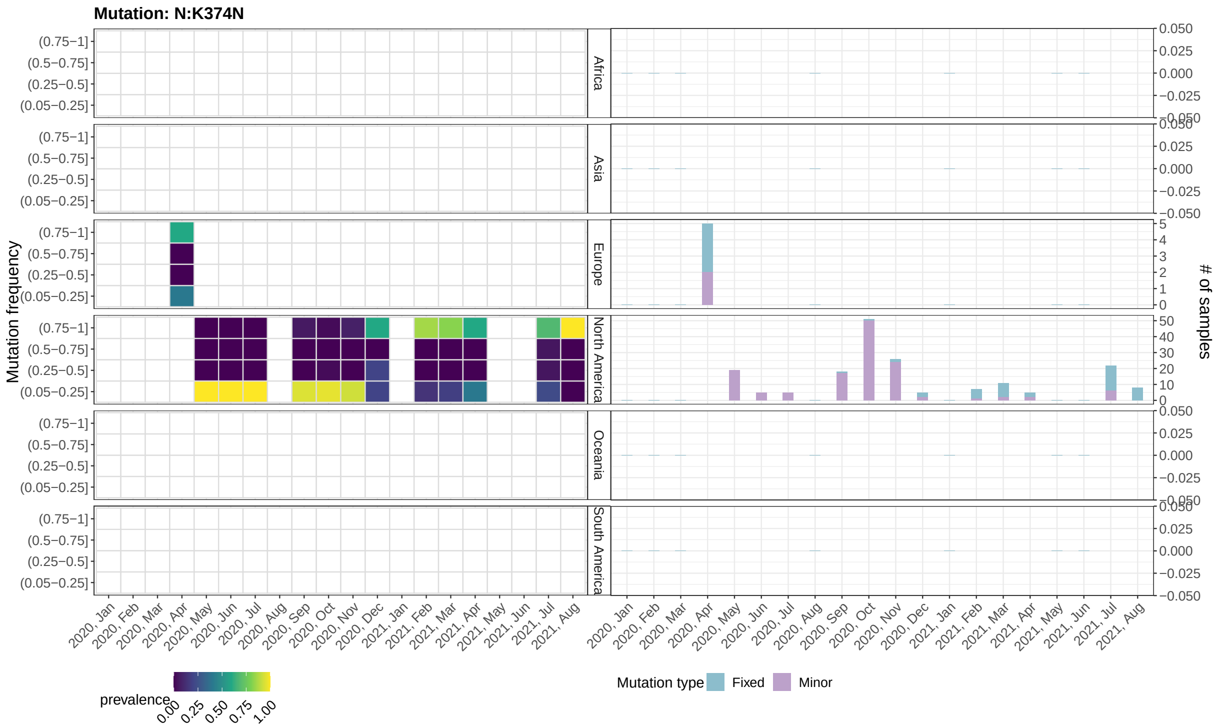


Figure 32: **Mutant frequency and prevalence variation in time of mutation: N:K374N.** Please refer to the caption of Supplementary Figure 3

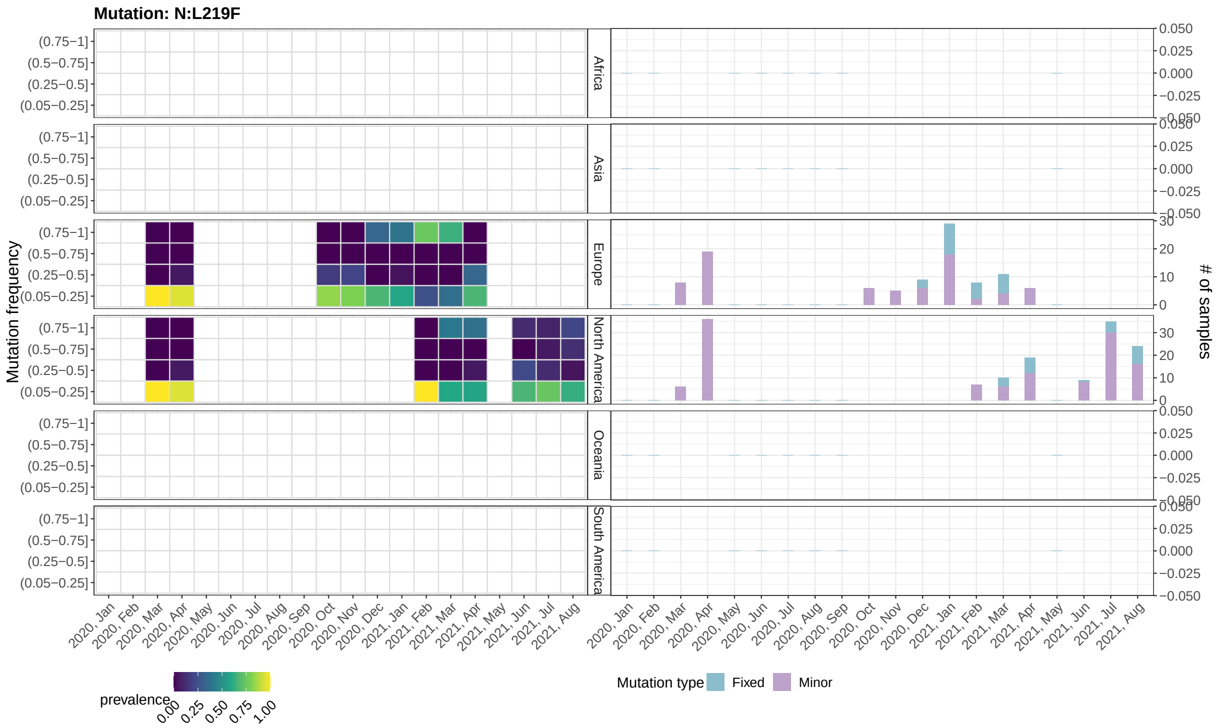


Figure 33: **Mutant frequency and prevalence variation in time of mutation: N:L219F.** Please refer to the caption of Supplementary Figure 3

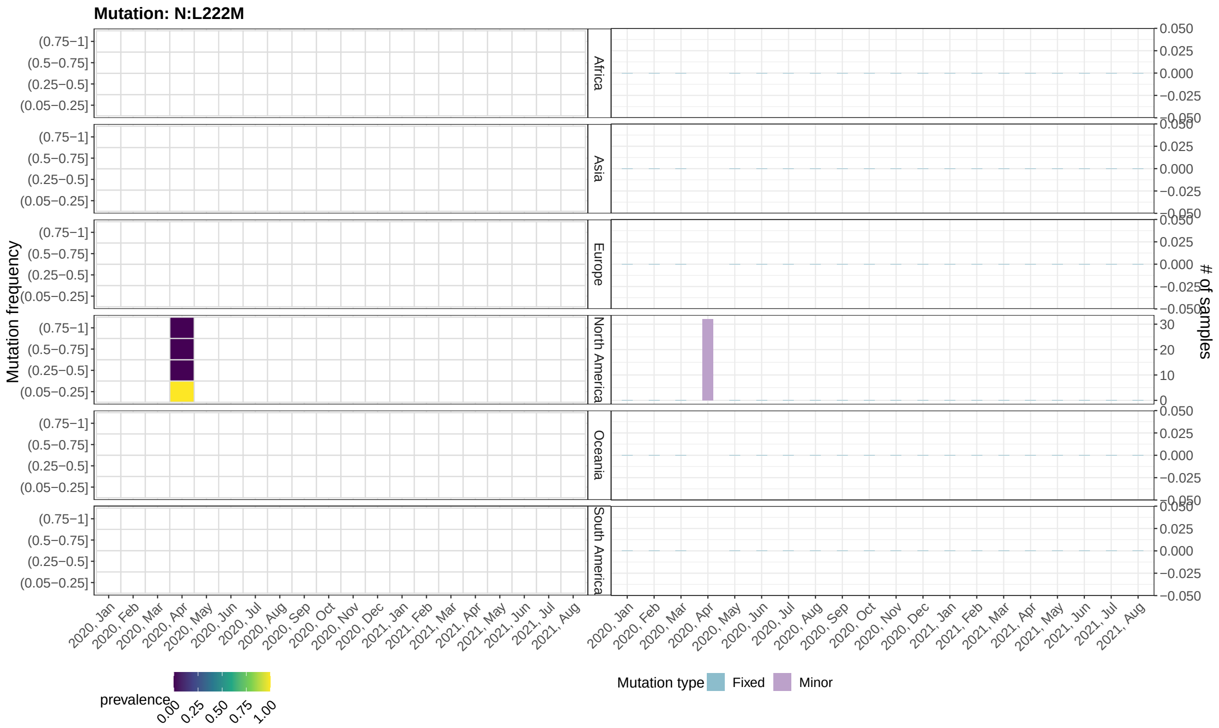


Figure 34: **Mutant frequency and prevalence variation in time of mutation: N:L222M.** Please refer to the caption of Supplementary Figure 3

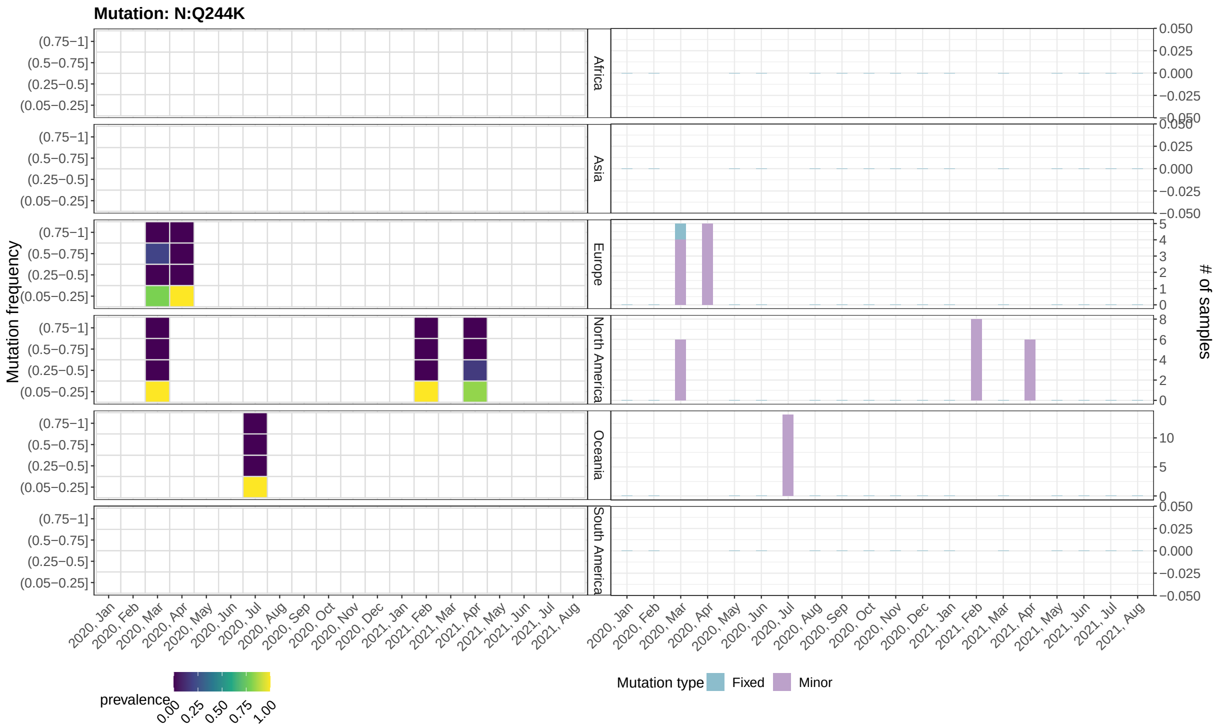


Figure 35: **Mutant frequency and prevalence variation in time of mutation: N:Q244K.** Please refer to the caption of Supplementary Figure 3

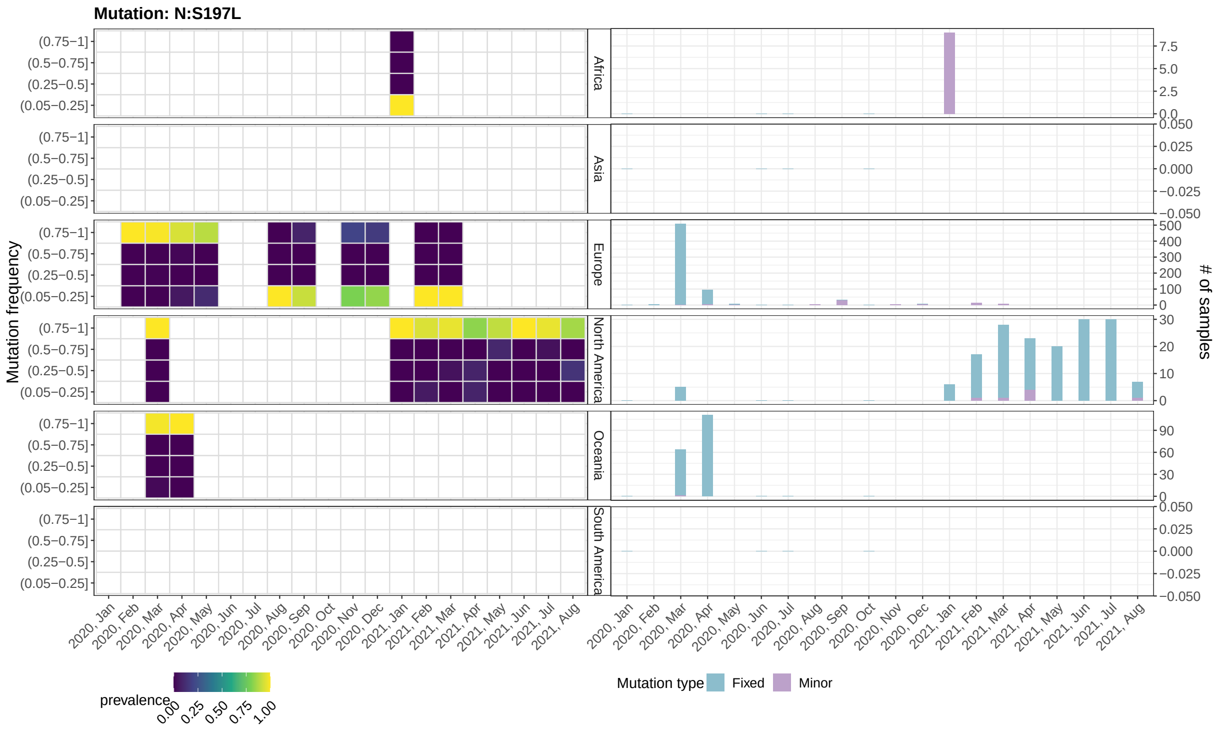


Figure 36: **Mutant frequency and prevalence variation in time of mutation: N:S197L.** Please refer to the caption of Supplementary Figure 3
